# Understanding Comorbidities in Hypermobile Ehlers-Danlos Syndrome: Could a Viral Infection Unmask the Disorder?

**DOI:** 10.1101/2025.10.22.25338573

**Authors:** Megan L. Pearson, Bryan J. Laraway, Ellen R. Elias, Ganna Bilousova, Melissa A. Haendel

## Abstract

Hypermobile Ehlers-Danlos Syndrome (hEDS) is a complex, underdiagnosed connective tissue disorder characterized by widespread symptoms affecting multiple organ systems. Recent clinical observations suggest that individuals with hEDS may be at increased risk for persistent symptoms following COVID-19, commonly referred to as Long COVID. Using data from over 19 million patients across the United States, we examined associations between hEDS, COVID-19 infection, Long COVID, and related chronic conditions.

We identified just over 25,000 individuals with hEDS and estimated a prevalence of approximately 1 in 800, which is higher than previously recognized. While rates of COVID-19 infection were similar between patients with hEDS and matched controls, patients with hEDS were significantly more likely to develop Long COVID. This risk was especially elevated among patients with hEDS with overlapping conditions commonly seen in post-viral syndromes, including autonomic dysfunction, immune dysregulation, and chronic fatigue. Specifically, individuals with postural orthostatic tachycardia, mast cell-related symptoms, or chronic fatigue syndrome had the highest rates of Long COVID.

Temporal diagnostic analyses revealed that many patients received an hEDS diagnosis only after a COVID-19 infection, suggesting that viral illness may exacerbate or reveal previously unrecognized symptoms. Patients with hEDS also exhibited higher odds of having additional risk factors for severe or prolonged illness, including chronic lung and autoimmune conditions, depression, and cerebrovascular disease.

These findings highlight a previously unrecognized vulnerability in patients with hEDS and underscore the need for greater clinical awareness of their heightened risk for persistent post-COVID illness. Improved screening, earlier diagnosis, and integrated care pathways are urgently needed to support this complex and underserved patient population.

**Author summary:** We studied patients with hypermobile Ehlers-Danlos Syndrome (hEDS), a connective tissue condition that affects joints, skin, and many body systems. This condition is often misunderstood or overlooked, leaving many people undiagnosed. During the COVID-19 pandemic, people with hEDS appeared to experience more long-term symptoms after infection, a condition often called Long COVID. Here, we analyzed the health records of millions of patients in the United States to better understand post-viral outcomes.

Patients with hEDS were more likely to be diagnosed with Long COVID compared to similar patients without hEDS. This was especially true for those who also had conditions such as chronic fatigue, immune conditions, or issues with heart rate and blood pressure regulation. In many cases, people were diagnosed with hEDS for the first time only after they had COVID-19, suggesting the virus may worsen or reveal symptoms that had been previously missed.

Our findings show that hEDS may be more common than previously thought and that such patients face higher risks after COVID-19. Greater awareness and earlier recognition of hEDS could improve care for many patients with complex, long-lasting symptoms

## Introduction

Hypermobile Ehlers-Danlos Syndrome (hEDS) is the most common subtype of the Ehlers-Danlos Syndromes (EDS), a group of inherited connective tissue disorders caused by pathogenic variants affecting extracellular matrix components and collagens[1–3]. Unlike the other 13 EDS subtypes, hEDS is the only subtype with no known molecular marker and is diagnosed clinically based on skin hyper extensibility, generalized joint hypermobility, systemic features, family history, and exclusion of other conditions[2,4]. hEDS is highly heterogeneous, with multisystem involvement that can include musculoskeletal pain, gastrointestinal dysmotility, autonomic dysfunction, and neurocognitive symptoms[5–11]. **Figure 1** highlights the range of symptoms and body systems that can be affected in hEDS, which vary widely across individuals in severity and combination[11–13].

**Figure 1.**
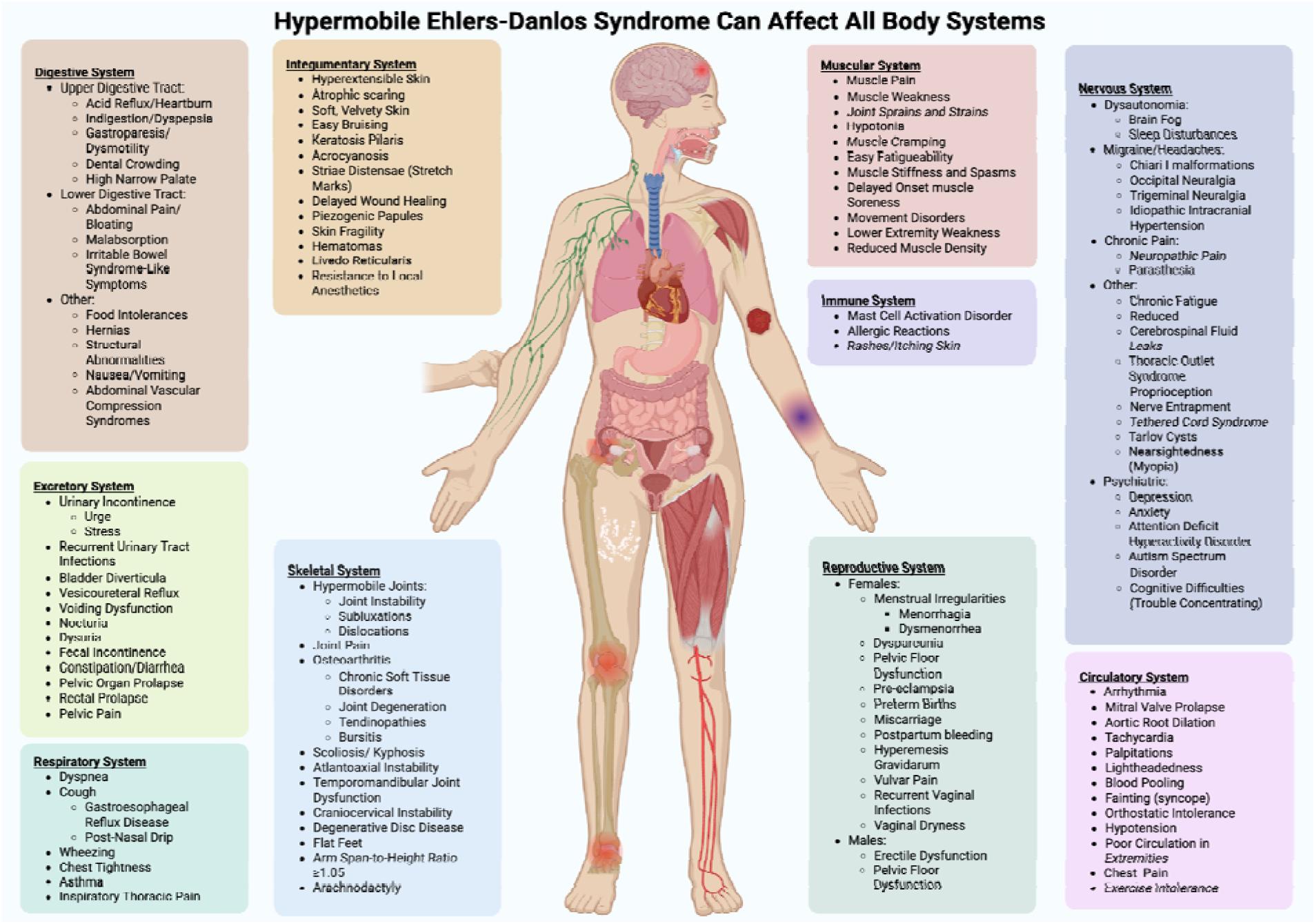
hEDS symptoms can affect all body systems. Patients with hEDS can exhibit manifestations in nearly all body systems making this complex disorder challenging to characterize and diagnose. Shown is an inclusive list of hEDS manifestations and frequently co-occurring conditions. The multisystem involvement without an apparent unifying cause frequently leads to diagnostic delays, as the underlying connective tissue disorder is not readily recognized. Created in BioRender. https://BioRender.com/8zctb43

This complexity contributes to frequent misdiagnosis or diagnostic delays. hEDS symptoms commonly overlap with better-recognized disorders such as fibromyalgia, irritable bowel syndrome, and psychiatric or somatic symptom disorders[6–8,14,15]. Autonomic features, including dizziness, syncope, and tachycardia, may meet criteria for postural orthostatic tachycardia syndrome (POTS)[9,10], while neurocognitive and affective symptoms are often attributed to anxiety or ADHD. The frequent co-occurrence in patients with hEDS of mast cell activation syndrome (MCAS), a poorly defined disorder marked by episodic immunological hypersensitivity and gastrointestinal symptoms, adds additional diagnostic ambiguity[16–19]. While patients may present with POTS and MCAS, this does not mean they automatically have a diagnosis of hEDS, as these can be co-occurring but are separate conditions. We hypothesize that hEDS is not a single disorder caused by a single gene mutation, but rather multiple disorders caused by many different genetic variants, which together give rise to the variable symptoms exhibited by patients with hEDS.

Joint hypermobility, a hallmark of hEDS, often diminishes with age or injury, making recognition more difficult over time[4,20–22]. Many systemic manifestations emerge or intensify after key life events such as puberty, pregnancy, or trauma, transitions often accompanied by hormonal shifts. Notably, approximately 80% of patients with hEDS are female, and growing evidence suggests a link between hEDS manifestations and hormonal influences, particularly estrogen[23]. These hormonal factors may modulate connective tissue integrity and symptom expression, contributing to sex-based differences in prevalence and clinical presentation. Without visible hypermobility or a known genetic marker, clinicians may not suspect a connective tissue disorder. Diagnosis is often delayed by years, with patients reporting prolonged medical odysseys involving multiple specialists and inconsistent care[2,24–26]. Given this diagnostic complexity, the actual prevalence of hEDS is likely underestimated.

Recent reports suggest patients with hEDS may also be at increased risk for post-acute sequelae of SARS-CoV-2 infection (COVID-19), commonly referred to as Long COVID [27–30]. This overlap may be driven by shared pathophysiologic features, including immune dysregulation (e.g., MCAS, autoimmunity), chronic inflammation, and dysautonomia (e.g., POTS). Structural tissue fragility may further predispose patients with hEDS to respiratory and vascular complications during and after COVID-19. Comorbidities common in hEDS, such as Myalgic Encephalomyelitis/Chronic Fatigue Syndrome (ME/CFS), chronic pain, and gastrointestinal dysfunction, also align with known Long COVID risk factors. Anecdotal and clinical observations suggest that SARS-CoV-2 infection may exacerbate or unmask underlying hEDS symptoms, triggering a new diagnosis. We aimed to determine whether a viral infection unmasks hEDS symptoms in some patients or whether patients are diagnosed based on classical ascertainment bias, whereby increased health care utilization leads to a higher likelihood of diagnosis.

Despite plausible biological and clinical connections, there is a lack of large-scale, population-level studies evaluating the relationship between hEDS and Long COVID. This study uses electronic health record (EHR) data from the National Clinical Cohort Collaborative (N3C) COVID-19 Enclave to examine the burden of COVID-19 and Long COVID among patients with hEDS, characterize key comorbidities, and assess whether viral infection may act as a diagnostic inflection point for previously unrecognized hEDS.

## Methods

### Data source: National Clinical Cohort Collaborative (N3C) COVID-19 enclave

This study utilized Level 2 de-identified data, including date-shifted event timestamps, from the N3C COVID-19 Enclave, a centralized, harmonized repository of EHRs contributed by more than 80 U.S. clinical institutions [30] (Data Use Request: RP-582482). All patient dates are uniformly shifted by up to ±180 days within each site to protect patient privacy. As a result, temporal relationships within each patient record are preserved, but cross-patient calendar dates are not comparable.

The N3C Enclave was established in April 2020 to facilitate national COVID-19 research. It includes three primary patient groups: (1) individuals tested for SARS-CoV-2; (2) those diagnosed with COVID-19 via laboratory confirmation or clinical coding; and (3) comparison patients with at least one encounter after January 1, 2020, but no record of COVID-19 infection or exposure [31]. Retrospective data from January 1, 2018, allow evaluation of pre-pandemic health status.

As of the December 2025 data release, the N3C contained over 19.8 million unique patient records. All data are harmonized to the Observational Medical Outcomes Partnership (OMOP) Common Data Model, enabling standardized querying across demographics, diagnoses, procedures, medications, labs, and clinical observations. Diagnoses from ICD-9/10-CM codes are mapped to OMOP concept IDs and stored in the condition occurrence table. Concept IDs used for this study are listed in Supplementary Table 1.

### Phenotype definitions and COVID-19/Long COVID classification

We leveraged the N3C Logic Liaison Table[31] to define COVID-19 cases, Long COVID, and related risk conditions[31]. This curated tool maps clinical phenotypes to OMOP concept sets using standardized inclusion/exclusion criteria. Supplementary Table 2 summarizes phenotypes used in this analysis. Long COVID codes were inconsistently used across time and institutions; prior to October 2021, B94.8 and non-specific codes were commonly used. As a result, both Long COVID and COVID-19 may be underrepresented in structured data.

### Cohort definition: hEDS identification

To identify patients with hEDS, we queried the N3C Data Enclave using OMOP concept IDs mapped to diagnostic codes for hEDS (**Figure 2**). In addition to patients explicitly diagnosed with hEDS, we also identified individuals coded with classical EDS (cEDS), vascular EDS (vEDS), and unspecified EDS. Because cEDS and vEDS are the next most common EDS subtypes after hEDS and have distinct ICD-to-OMOP mappings, we excluded individuals with these diagnoses to avoid misclassification. Given that 80–90% of all EDS cases are believed to be hEDS, we included patients with the unspecified EDS code as suspected hEDS cases. We identified 7,955 patients with a specific hEDS diagnosis and 24,834 patients with the unspecified EDS code. To ensure data completeness, we excluded patients lacking demographic information, including age, sex, race, and ethnicity. After filtering, the final diagnosed hEDS group included 7,786 patients, and the suspected hEDS group included 24,031 patients. These two groups were combined to create a final hEDS cohort of 26,108 individuals. To note, 5,709 patients had both unspecified EDS and hEDS coded in their charts. To confirm that cEDS and vEDS cases were not inadvertently included in the final cohort, we separately identified patients with those diagnoses and retained only those with complete demographic data, 562 cEDS and 600 vEDS patients respectively. The final hEDS cohort thus consisted of 25,192 patients. Also of note, only 216 patients had both a diagnosis of cEDS or vEDS and an unspecified EDS diagnosis, so only those patients were removed. At the time of analysis, the total N3C population included approximately 19.8 million patients.

**Figure 2:**
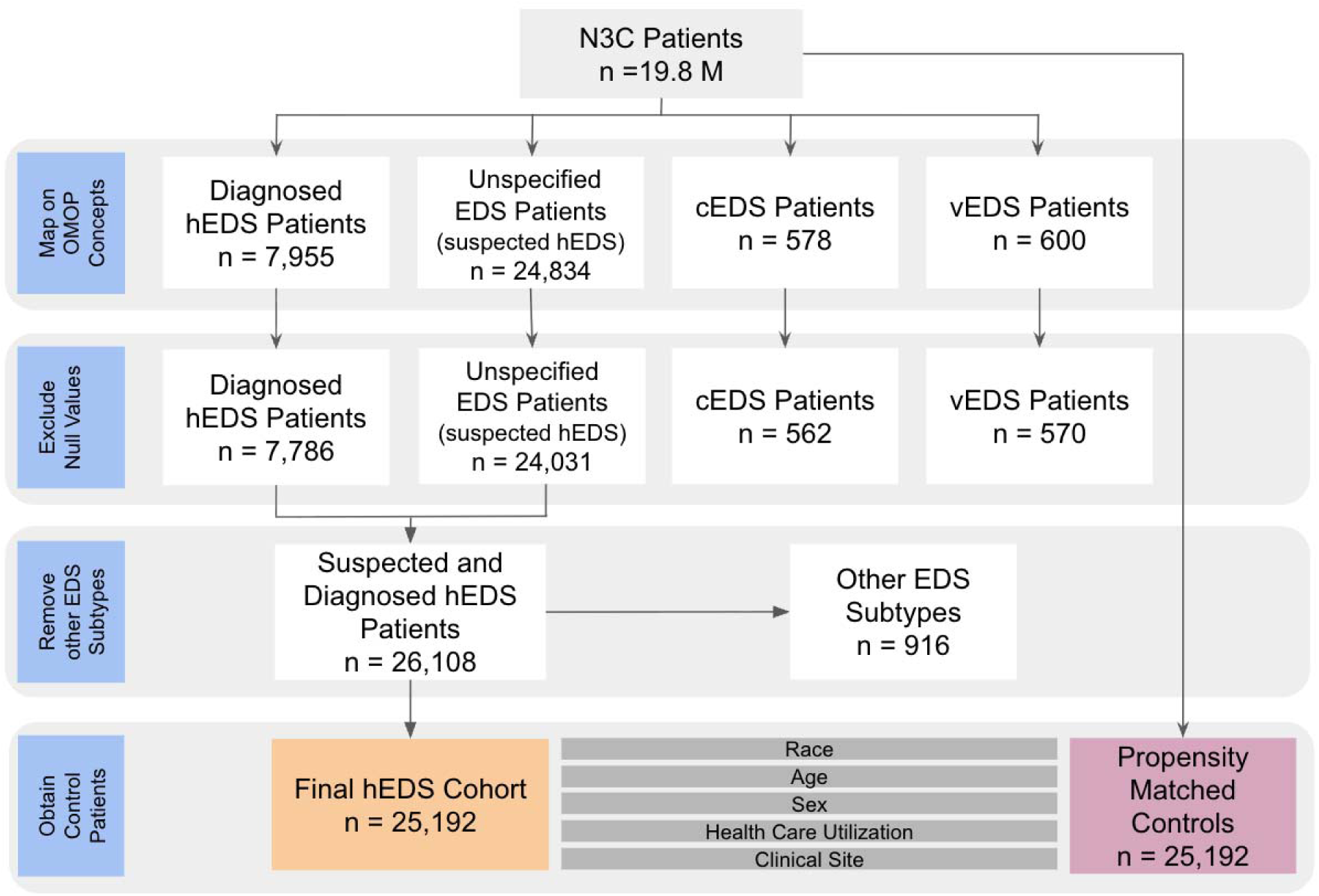
Cohort construction. Patients in the N3C database with a diagnostic code for hEDS were used to construct the cohort. Diagnosed hEDS cases were identified using the specific condition ID for hEDS, while suspected cases were identified using the condition ID for EDS with unspecified subtype. Patients with missing values for sex, age, race, or ethnicity were excluded. To minimize inclusion of other EDS subtypes, individuals with diagnostic codes for classical (cEDS) or vascular EDS (vEDS) were also excluded, yielding a final cohort of 26,108 patients. Propensity matched controls for age, sex, race, ethnicity, healthcare utilization, and clinical site were obtained from the base N3C patient population of 19.8M.

### Comorbidity identification and diagnosis dating

We focused on three conditions frequently co-occurring with hEDS and Long COVID: MCAS, POTS, and ME/CFS. Patients were flagged as positive for each comorbidity if they had ≥1 recorded OMOP concept. For each condition, as well as for hEDS, COVID-19, and Long COVID, the initial diagnosis date was set as the earliest recorded condition start date for the relevant OMOP concept identifiers. Patients flagged as positive for a condition were required to have a non-null first diagnosis date for that condition; records that did not meet this criterion were excluded. Diagnosis dates were used to derive patient-level temporal relationships between hEDS and COVID-19, Long COVID, and comorbid diagnoses, including whether hEDS occurred before, within six months after, or more than six months after the comparator diagnosis. Patients may have multiple comorbidities to reflect real-world multimorbidity patterns.

All SQL, R, and Python workflows used for cohort construction, feature extraction, statistical modeling, and survival analyses were executed as numbered analysis nodes in the N3C Workbench; the complete, versioned codebase is publicly available and archived (Zenodo DOI: 10.5281/zenodo.18508715).

### Statistical Analyses

#### Prevalence Estimation

Prevalence rates were calculated to describe the burden of hEDS, comorbidities, COVID-19 infection, and Long COVID, within the study cohort. The denominator included all individuals in the N3C Enclave with at least one clinical encounter between 2018 and 2025 and complete demographic information. Cases were identified using OMOP concepts outlined in the cohort construction and comorbid conditions. Point prevalence was calculated as the proportion of individuals meeting case criteria at any time during the study window with 95 percent confidence intervals. To calculate a 1-in-X representation, we used the inverse of the point prevalence, with 95 percent confidence intervals.

#### Propensity Score Matching

To control for potential confounding by demographic, institutional differences, and healthcare utilization trends, we constructed a propensity score–matched control cohort using the MatchIt package in R [32]. One-to-one nearest-neighbor matching without replacement was performed based on age, sex, race, total visits, and data partner (i.e., contributing clinical site). Matching on data partners was included to mitigate potential bias arising from variability in diagnostic coding practices and population demographics across institutions. Matching on health care utilization was performed to assess true differences compared to ascertainment bias. Healthcare utilization was assessed at the person level using the OMOP visit occurrence table. To reduce bias from multiple encounters recorded on the same day for a single individual, visits were collapsed into a single per day visit, (similar to other approaches documented in Pfaff <u>et.al</u> [33]). Distinct visit dates were counted to derive the total number of healthcare encounters, with visit-type specific counts for inpatient, outpatient, and emergency department visits based on standard OMOP visit concept identifiers. The resulting control cohort comprised 25,192 individuals with no recorded diagnosis of EDS, including hEDS, cEDS, vEDS, or unspecified EDS subtypes. A table depicting covariate balance between cases and controls is shown in **Supplementary Table 3**.

#### Odds Ratios

We evaluated associations between hEDS and predetermined COVID-associated phenotypes[31] described above, using conditional logistic regression on propensity-matched data to account for the matched pairs. No additional covariates were included. For each category, we obtained an odds ratio (OR), 95% confidence interval (CI), and Benjamini-Hochberg false discovery rates (FDR) adjusted q-value. The total number of tests was equal to the number of phenotype categories evaluated in **Figures 3** and **7** (N=16, 36, respectively). We controlled the FDR at five percent and considered categories with q less than 0.05 as significant. The figures display the OR and 95 percent CI for each category along with the corresponding FDR q-value.

**Figure 3.**
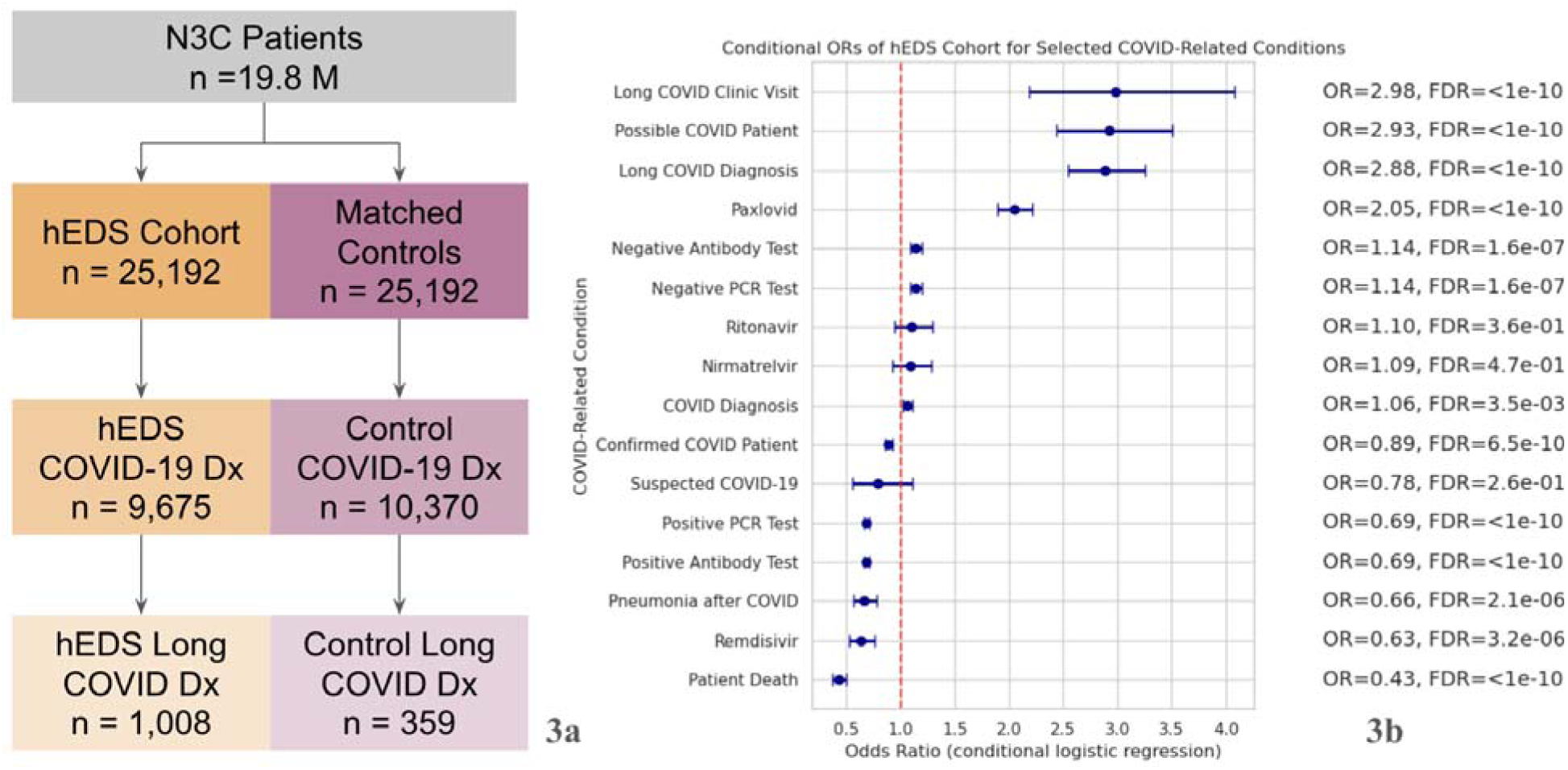
Patients with hEDS are more likely to develop Long COVID despite similar rates of COVID-19 infection 3a. Counts of COVID-19 cases (OMOP Concept ID: 37311061) and Long COVID diagnoses (OMOP Concept ID: 705076) are shown for the hEDS cohort (orange) and matched controls (pink). **3b.** Odds ratios (ORs) with 95% confidence intervals were calculated for COVID-19–related diagnoses and medications using phenotype categories defined in the N3C Enclave (see methods). Multiple testing was addressed using Benjamini–Hochberg false discovery rate correction; q values are shown in the figure. Patients with hEDS showed increased odds of Long COVID diagnoses and Long COVID clinic visits compared to controls.

#### Cumulative Incidence Analyses

To assess the temporal relationships between diagnoses of hEDS, COVID-19, Long COVID, and related comorbid conditions, we performed cumulative incidence analyses using Kaplan-Meier estimators. Analyses were conducted on a cohort of individuals with known hEDS diagnoses, comorbid symptoms, and COVID-19 confirmed infection history. Dates of initial diagnoses were extracted from clinical records for hEDS, COVID-19, Long COVID, MCAS, POTS, and ME/CFS. For each analysis, patients lacking a recorded diagnosis date for the index condition were excluded, and patients without the event of interest were censored at the end of their available follow-up. Due to Level 2 de-identification, all dates were shifted by up to ±180 days per patient per site, though within-patient temporal order was preserved. Cumulative incidence analyses focused on hEDS diagnosis following COVID-19 or Long COVID, restricted to patients with documented COVID-19 or Long COVID diagnoses. Patients with hEDS diagnosed prior to the index condition were excluded. Time to event was defined as the number of days from the first recorded COVID-19 or Long COVID diagnosis to the first hEDS diagnosis or censoring. Kaplan–Meier survival functions were inverted to estimate cumulative incidence and stratified by comorbidity status (MCAS, POTS, ME/CFS, or no comorbidities). Group-specific 95% confidence intervals were calculated and plotted. Pairwise differences between strata were assessed using log-rank tests **(Supplemental Table 6)**. Analyses were performed in Python using the lifelines package [34]. **Supplemental Figure 3** presents the cumulative incidence curves for all patients, including those without recorded COVID-19 or Long COVID diagnoses, to illustrate the full cohort.

#### Cox Regression (Post-Covid hEDS diagnostic unmasking analysis)

We conducted a Cox proportional hazards sensitivity analysis to evaluate factors associated with post-COVID documentation of hEDS shown in **Table 4**. Individuals with a documented hEDS diagnosis prior to SARS-CoV-2 infection were excluded to restrict the cohort to those at risk for post-COVID diagnosis. Time zero was defined as the date of the first documented COVID-19 infection. The event of interest was the first recorded hEDS diagnosis after the COVID-19 index date. Follow-up time was calculated in days from the index date to the hEDS diagnosis or administrative censoring at the latest available data cut. Indicators of comorbid conditions, including MCAS, POTS, and ME/CFS, were defined as binary variables reflecting documentation at any time during follow-up and were interpreted as phenotypic markers rather than time-ordered predictors. A composite indicator reflecting the absence of documented comorbidities was included as a descriptive reference phenotype. Cox models were fitted with L2 penalization (penalizer = 0.1) to ensure numerical stability. Hazard ratios (HRs) with 95% confidence intervals (CIs) are reported. Analyses were conducted in Python using the lifelines package [34].

#### Time Series Analysis

To evaluate temporal patterns in first hEDS diagnoses relative to COVID-19, we calculated monthly counts of incident diagnoses for each patient category: hEDS before COVID, hEDS after COVID, and hEDS never COVID. Monthly counts were normalized to the total number of clinical encounters per month, yielding diagnoses per 1,000 encounters. Months with fewer than 20 encounters were masked to comply with data suppression requirements. Trends were visualized using line plots showing both normalized and raw counts over time.

## Results

### hEDS is more prevalent than previously recorded

As shown in Table 1, we calculated the prevalence of hEDS using both hEDS and unspecified EDS diagnoses together as one cohort and hEDS diagnoses alone as the other cohort. Using the full hEDS cohort described in the Methods, we calculated a prevalence of 0.127% or 1 in 788 (95% CI, 1 in 778, 1 in 798) patients within the N3C Enclave. This rate is substantially higher than current estimates (∼1 in 3,100) [35]but consistent with prior findings in population-based studies, such as those conducted in Wales [24]. Additionally, we found the prevalence for patients with a specified hEDS diagnosis to be 1 in 2,549 (95% CI 1 in 2,494, 1 in 2,607), still higher than previously reported.

**Table 1:** hEDS Prevalence Estimation. We estimated the point prevalence of hEDS within our database, and recorded a 95 percent confidence interval (CI) by using the number of cases (25,192 and 7,786) with full demographic information. To calculate the 1 in X we took the inverse of the prevalence calculated.

| Cohort (N) | Prevalence (95% CI) | 1 in X (95% CI) |
| --- | --- | --- |
| hEDS + Unspecified EDS (25,192) | 0.127% (0.125% - 0.129%) | 1 in 788 (1 in 778 - 1 in 798) |
| Only hEDS (7,786) | 0.039% (0.038% - 0.040%) | 1 in 2,549 (1 in 2,494 - 1 in 2,607) |

### hEDS cohort demographics: diagnosed patients are mostly white, Non-Hispanic females aged 25-45

**Table 2** presents the demographic characteristics of the hEDS cohort. The majority of diagnosed patients were female (84.21%), White (88.33%), and Non-Hispanic (90.53%). Nearly half (48.97%) were aged 26–45. These findings are consistent with previous reports on hEDS epidemiology [24]. A matched control cohort had a similar demographic distribution based on propensity matching, as shown in Supplementary Table 3.

**Table 2.** hEDS Cohort Demographics. hEDS patient population in N3C is majority white, non-hispanic, females, with an age range of 25-45 primarily. American Indian or Alaska Native = AI/AN. Native Hawaiian or Other Pacific Islander = NH/PI.

| hEDS Cohort Demographics |  | n = 25,192 (%) |
| --- | --- | --- |
| **Sex** |  |  |
|  | Female | 21,180 (84.21) |
|  | Male | 3,214 (12.78) |
|  | Other/Unknown | 758 (3.01) |
| **Ethnicity** |  |  |
|  | Non-Hispanic | 22,770 (90.53) |
|  | Hispanic | 1,381 (5.49) |
|  | Other/Unknown | 1,001 (3.98) |
| **Race** |  |  |
|  | White | 22,218 (88.33) |
|  | Other/Unknown | 1,778 (7.07) |
|  | Black | 686 (2.73) |
|  | Asian | 257 (1.02) |
|  | AI/AN | 175 (0.7) |
|  | NH/PI | 38 (0.15) |
| **Age** |  |  |
|  | 0-18 | 2,009 (7.99) |
|  | 18-25 | 3,857 (15.33) |
|  | 26-35 | 6,975 (27.73) |
|  | 36-45 | 5,342 (21.24) |
|  | 46-55 | 3,496 (13.9) |
|  | 56-65 | 2,040 (8.11) |
|  | 66-75 | 1,052 (4.18) |
|  | 76-100 | 318 (1.51) |

### Patients with hEDS have higher Long COVID rates despite similar SARS-CoV-2 infection rates

To assess the relationship between hEDS and COVID-19 outcomes, we first compared rates of documented SARS-CoV-2 infection and Long COVID between patients with hEDS and propensity-matched controls, shown in **Figure 3a**. Using the OMOP concept for confirmed COVID-19 infection (ID:37311061), we found that 9,675 (38.4%) individuals in the hEDS cohort had evidence of COVID-19 in their EHR, compared to 10,370 (41.2%) in the matched control group, indicating similar infection rates between the two populations. However, rates of Long COVID were markedly different. Using the OMOP identifier for Long COVID (ID:705076), 1,008 (4.0%) of patients with hEDS had documentation of Long COVID, compared to only 359 (1.4%) matched controls, representing a 2.8-fold higher proportion of Long COVID cases than controls.

We then assessed COVID-19-related outcomes and treatments across both groups (**Figure 3b**). All odds ratios were evaluated using conditional logistic regression, with multiple testing controlled using Benjamini–Hochberg false discovery rate correction. Odds ratio (OR) estimations confirmed that patients with hEDS had substantially higher odds of receiving a Long COVID diagnosis (OR = 2.88, 95% CI: 2.55-3.26) and attending a Long COVID specialty clinic (OR = 2.98, 95% CI: 2.18-4.08). While the odds of having a confirmed COVID-19 diagnosis were not elevated in patients with hEDS (OR = 0.89, 95% CI: 0.86-0.92; q 6.4 e^-10^), the odds of being classified as a “possible COVID-19 case” based on symptoms or testing history were substantially higher (OR = 2.93, 95% CI: 2.44-3.51).

Treatment-related data revealed increased odds for the use of COVID antiviral therapies among patients with hEDS. Specifically, the odds of receiving Paxlovid were over 2 times higher in patients with hEDS than controls (OR = 2.05, 95% CI:1.89-2.21). Remdesivir use did not differ significantly between groups (OR = 0.63, 95% CI: 0.53-0.76). In contrast, testing-related variables, such as the presence of a negative PCR or antibody test, showed no difference between groups, nor did outcomes like pneumonia or mortality following COVID-19 infection. All the associations described above remain statistically significant after the FDR correction.

These results suggest that while patients with hEDS are not more likely to contract COVID-19 than matched controls, they are substantially more likely to develop post-acute sequelae, to receive targeted antiviral therapies, and to seek specialized care for Long COVID.

### Timing of hEDS diagnosis relative to COVID-19 infection

To understand how COVID infection rates related to hEDS diagnosis in our cohort, we grouped the hEDS patients into timing categories: those diagnosed before COVID-19, more than 6 months after COVID-19, or within 6 months after COVID-19, as shown in **Table 3**. While most hEDS patients had a diagnosis prior to a COVID-19 infection (63%), a substantial fraction were diagnosed after COVID-19 (27%). In this group, COVID-19 may act as a trigger for clinical recognition. The 10% of patients diagnosed with hEDS within six months of a COVID-19 infection likely represent a subgroup with increased diagnostic acceleration, spikes in healthcare utilization, and Long COVID workups.

**Table 3.** Diagnostic Timing of hEDS Diagnosis and COVID-19 Infection. . Among 9,445 individuals with both hEDS and COVID-19 infection, 63% of them had a documented hEDS diagnosis prior to COVID-19 infection, while 27% received their first hEDS diagnosis after infection, including 10% within six months. *This number differs from the total calculated COVID positive hEDS patients because some are missing infection dates and therefore not captured in this calculation.

| Timing Group | N= 9,445* (%) |
| --- | --- |
| hEDS within 6 months after COVID-19 | 933 (10) |
| hEDS more than 6 months after COVID-19 | 2547 (27) |
| hEDS before COVID-19 | 5965 (63) |

### Temporal trends in first hEDS diagnoses across the COVID-19 pandemic

Next, we wanted to assess temporal trends of hEDS diagnoses in relation to COVID infection and how this changed over time during the COVID-19 pandemic. In raw counts, distinct temporal patterns were observed across COVID-19 exposure groups **(Figure 4a)**. The first recorded hEDS diagnoses among patients whose diagnosis occurred before COVID-19 infection declined over time, while diagnoses following COVID-19 infection increased. Patients without documented COVID-19 consistently accounted for the largest number of hEDS diagnoses, with counts remaining relatively stable early in the study period and gradually declining thereafter. When stratified by Long COVID status, similar directional patterns were observed, although absolute diagnosis counts were substantially lower **(Figure 4b)**.

**Figure 4.**
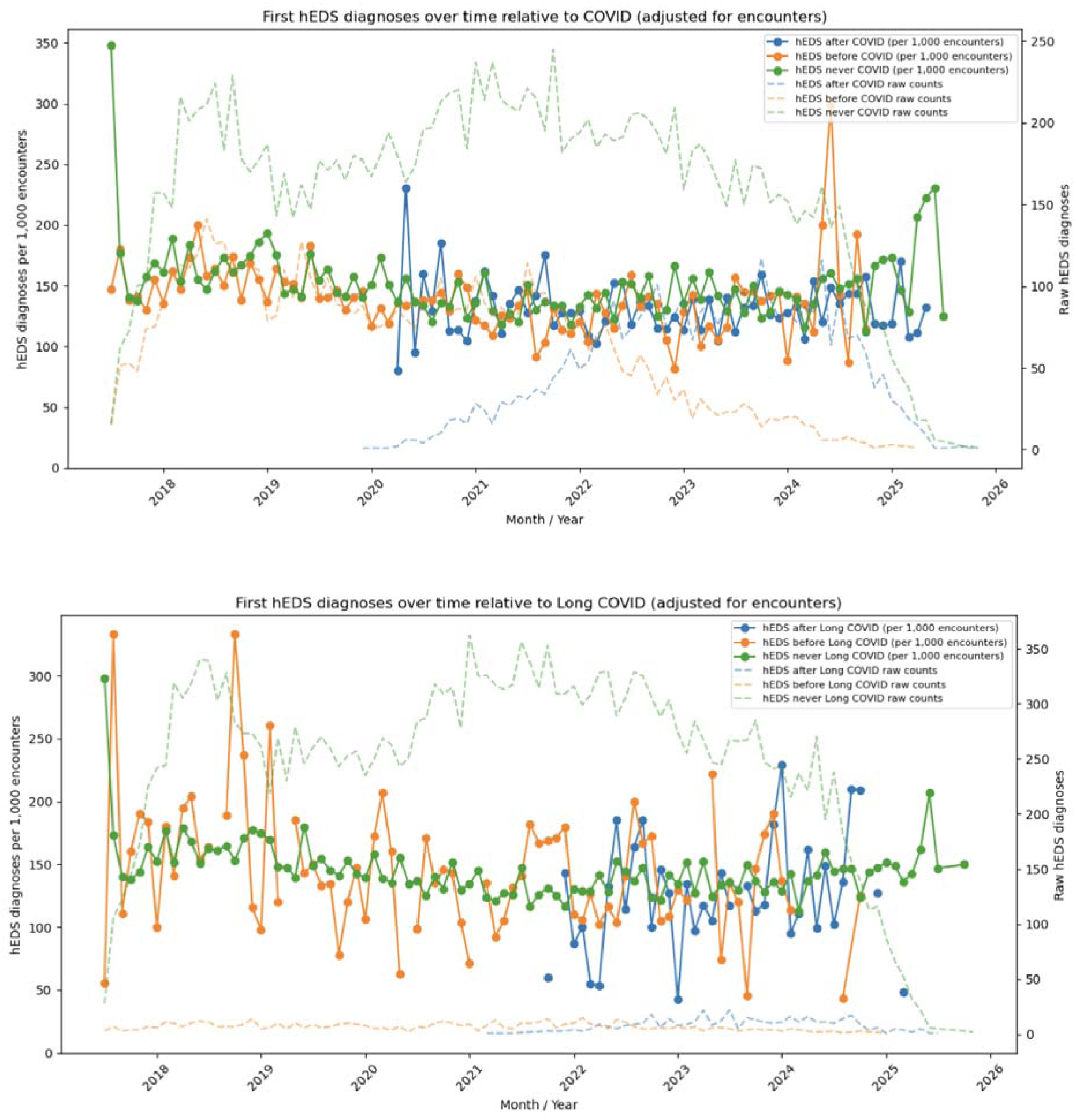
Monthly first hEDS diagnoses over time (adjusted per encounters + raw counts). To assess diagnostic rates of hEDS relative to COVID-19 infection, we assessed both raw counts and per encounter frequency (per 1,000 encounters) over time. **4a** shows the first hEDS diagnosis relative to COVID-19 infection (after, before, never). **4b** shows the first hEDS diagnosis relative to the development of Long COVID (after, before, never). When adjusted for encounter frequency, there is no change in hEDS diagnosis rate based on COVID-19 infection status or Long Covid development.

To determine whether these temporal shifts reflected typical ascertainment bias driven by increased healthcare utilization during the pandemic, hEDS diagnoses were normalized to encounter frequency. After adjustment, hEDS diagnosis rates per 1,000 encounters remained consistent over time and showed no meaningful differences by COVID-19 timing or Long COVID status. Despite varying raw count trajectories across groups, encounter-adjusted diagnosis rates did not increase over time following COVID-19 or Long COVID.

Under a classical ascertainment bias model, increased healthcare contact after infection would likely lead to higher diagnosis rates per encounter over time. The lack of such an increase suggests that the observed shift in raw hEDS diagnosis counts across COVID-19 exposure groups is due to changes in healthcare utilization rather than an increase in diagnostic success per encounter, arguing against classical ascertainment bias as the main cause of these trends.

### Patients with hEDS are more likely to receive a diagnosis after COVID-19 and Long COVID when comorbid conditions are present

Next, we aimed to assess whether patients with immune (MCAS), autonomic (POTS), or fatigue-related (ME/CFS) comorbidities had significant effects on the timing of hEDS diagnosis when COVID-19 infection or Long COVID developed. Among patients with hEDS who experienced COVID-19 or Long COVID (N=9,675 and 1,008 respectively), the likelihood of receiving an hEDS diagnosis was highest in those with autonomic dysfunction (POTS) or immune dysregulation (MCAS), followed by those with chronic fatigue (ME/CFS). Kaplan-Meier curves **(Figure 5a–5b)** show that patients with POTS or MCAS reached a considerably higher cumulative incidence of hEDS diagnosis after infection, whereas ME/CFS was associated with a more moderate increase. Log-rank tests showed significant differences between ME/CFS and all other groups (p≤0.001), while differences between POTS and MCAS were not statistically significant (p>0.1). Detailed day-specific cumulative incidences and all pairwise log-rank p-values are provided in **Supplemental Table 6**. Patients with hEDS but without these comorbidities had substantially lower cumulative incidence. These patterns suggest that overlapping symptom syndromes contribute to diagnosis risk, but POTS and MCAS appear to be the strongest factors driving faster or more frequent hEDS identification. These findings support the hypothesis that a viral infection might act as a physiological stressor, unmasking latent connective tissue dysfunction in genetically susceptible individuals.

**Figure 5.**
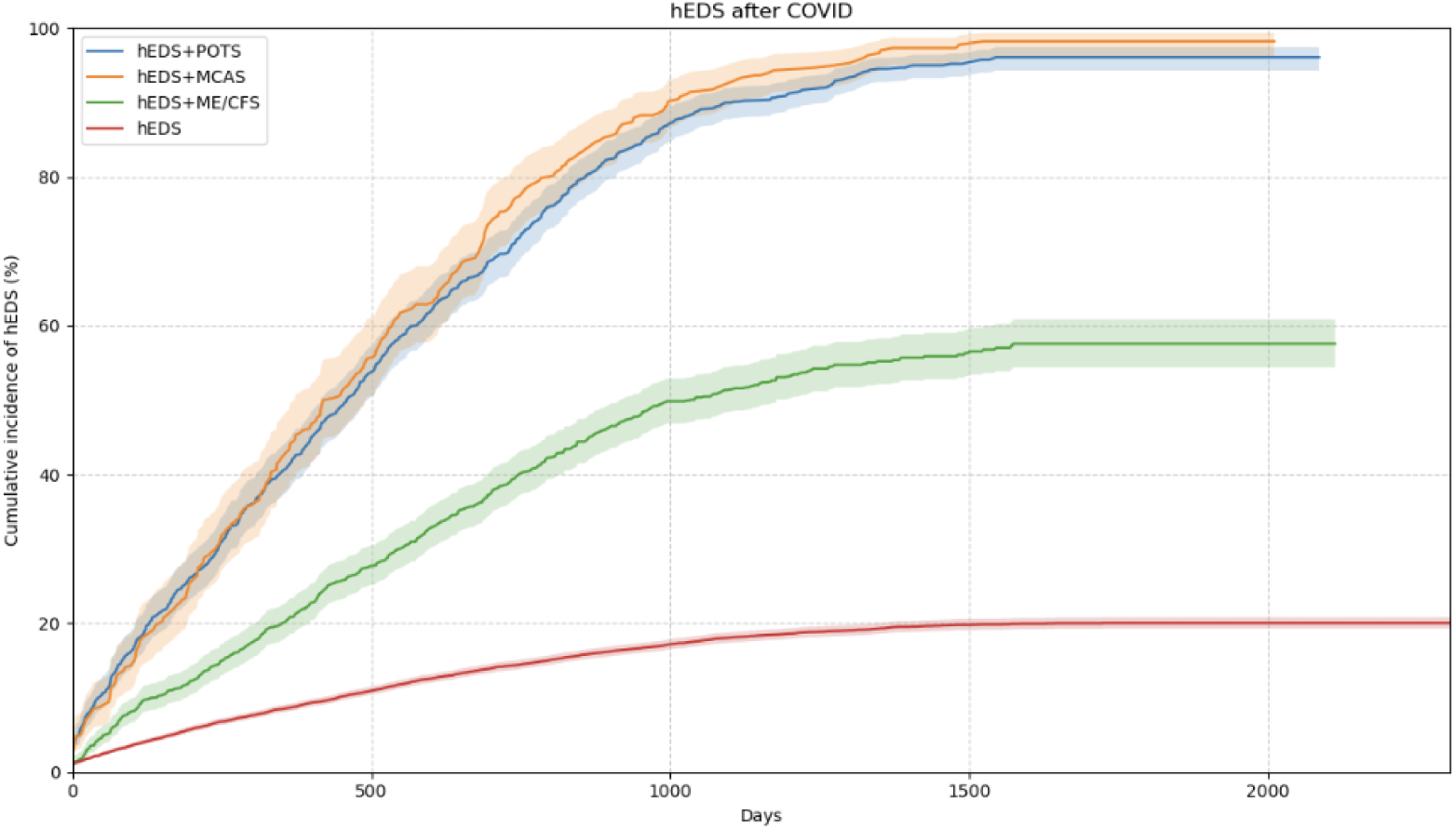
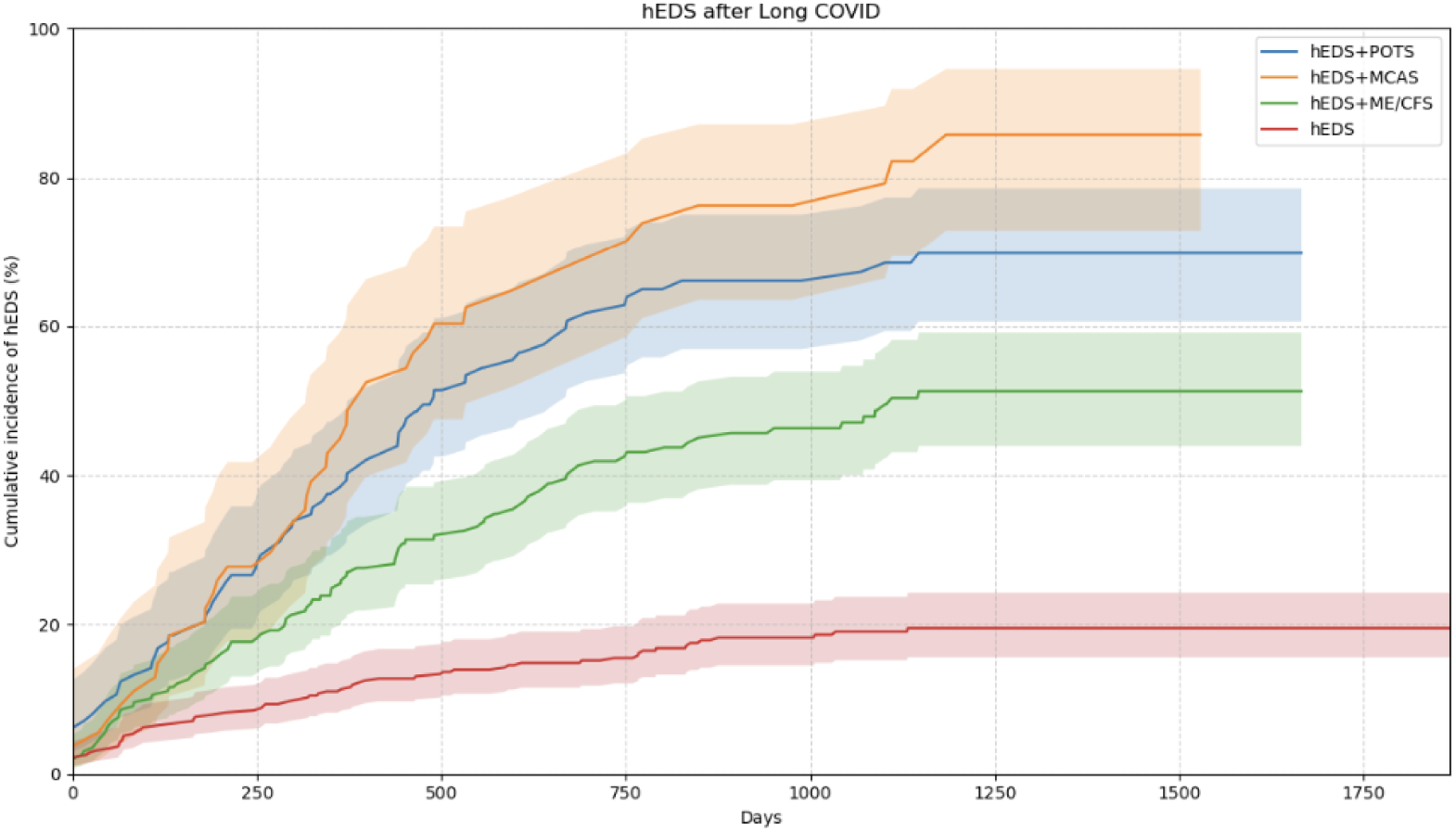
Cumulative incidence plots show that patients with hEDS have a higher likelihood of receiving a diagnosis after an acute post-viral infection with complications when comorbidities are present. Cumulative incidence plots demonstrate that patients with hEDS face an increased chance of receiving a diagnosis following an acute post-viral infection and experience a greater risk of complications when comorbid conditions are present. Panels show the cumulative incidence of receiving an hEDS diagnosis after COVID-19 infection **(5a)** or Long COVID (**5b)**. Time is measured in days from the index event, COVID-19 diagnosis, and the cumulative incidence was estimated using the Kaplan-Meier method. Shaded regions represent 95% confidence intervals.

To statistically evaluate these findings, we also performed a Cox proportional hazards sensitivity analysis restricted to individuals without hEDS prior to COVID-19 infection. We found that documented POTS and MCAS were associated with significantly higher hazards of subsequent hEDS diagnosis (HR 3.28, 95% CI 2.65-3.28; HR 1.82, 95% CI 1.60-2.33, respectively). ME/CFS was not independently associated with post-COVID hEDS diagnosis (HR 1.08, 95% CI 0.97 - 1.19). Absence of documented comorbidities correlated with a lower hazard of hEDS diagnosis during follow-up (HR 0.43, 95% CI 0.39 - 0.48). These results align with the cumulative incidence patterns shown by the Kaplan-Meier curves, indicating that autonomic dysfunction and immune dysregulation are the primary factors driving accelerated hEDS diagnosis following COVID-19, while chronic fatigue alone does not significantly increase diagnostic likelihood. Together, these findings suggest that overlapping symptom syndromes, especially POTS and MCAS, may reveal latent connective tissue disorders in the post-infectious setting.

**Table 4.**
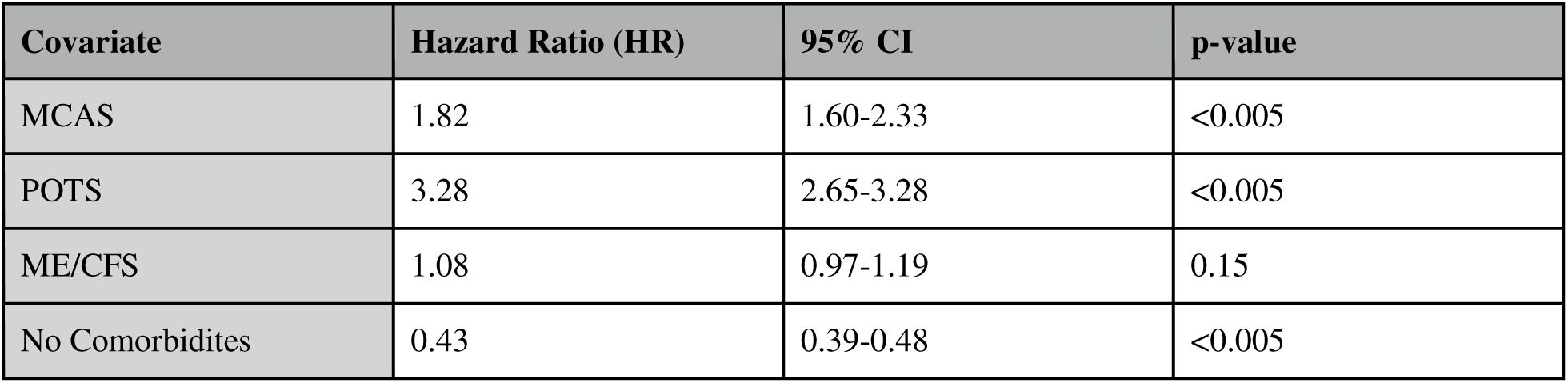
Cox Proportional Hazards Sensitivity Analysis. Hazard ratios were estimated using Cox proportional hazards models adjusted for all listed covariates. HR > 1 indicates higher hazard of post-COVID hEDS diagnosis.

### Long COVID is more common in patients with hEDS who also have comorbid MCAS, POTS, and ME/CFS

To explore the connection between common comorbidities and Long COVID in patients with hEDS, we examined the prevalence and co-occurrence of MCAS, POTS, and ME/CFS among hEDS patients who developed Long COVID and compared these patterns to their overall occurrence in the hEDS population (**Figure 6, Supplementary Table 5**). Among hEDS patients who developed Long COVID, POTS was the most prevalent comorbidity (42.7%), followed by ME/CFS (42.0%) and MCAS (25.3%) (**Figure 6a**). Multimorbidity was common in this group. MCAS and POTS co-occurred in 19.3% of Long COVID cases, MCAS and ME/CFS in 14.1%, and POTS and ME/CFS in 21.8%. Notably, 11.9% of hEDS patients with Long COVID had diagnoses of all three conditions. When compared to the overall prevalence of these comorbidities in the entire hEDS cohort (**Figure 6b**), patients with Long COVID showed a significant increase in both individual comorbidities and multi-system overlap. Although MCAS, POTS, and ME/CFS were present in the broader hEDS population, their co-occurrence was substantially less frequent outside of the Long COVID subgroup.

**Figure 6.**
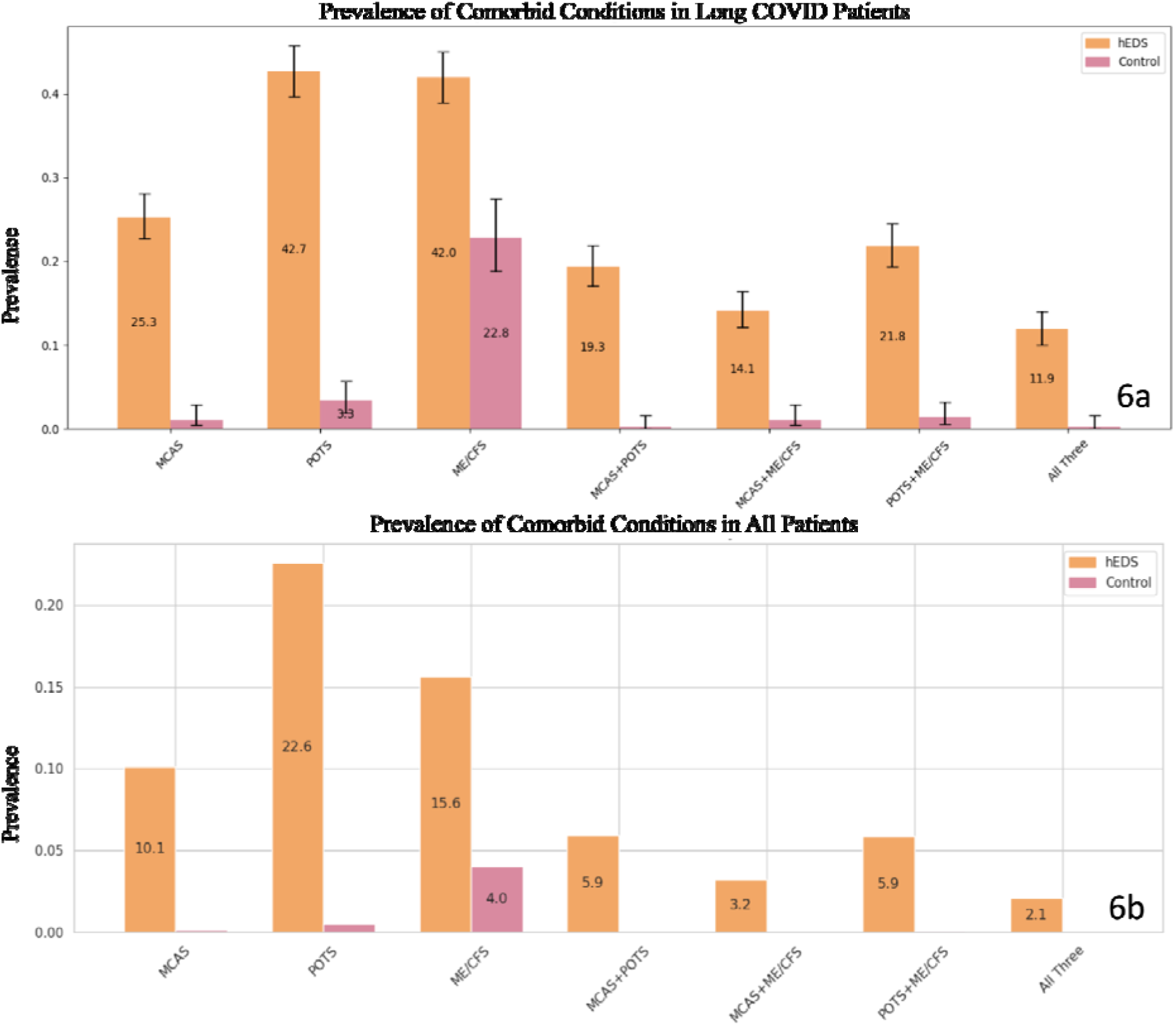
Higher prevalence of comorbid conditions in patients with hEDS who develop Long COVID. Prevalence rates are reported for commonly recognized comorbid conditions in hEDS (25,152) and in those developing Long COVID (1,008) as well as controls (25,152) and (359), respectively. Prevalence rates for condition combinations are also shown. To protect patient confidentiality, counts fewer than 20 are listed as <20 or left blank. The prevalence of comorbid conditions is higher in patients with hEDS who develop Long COVID, indicating that having these comorbidities may be risk factors for developing Long COVID.

We also assessed these patterns among COVID-19–positive hEDS patients regardless of Long COVID status and found similar, though less significant, enrichment of comorbidities (**Supplementary Figure 1**). This suggests that a higher burden of multisystem comorbidities is more strongly linked to the progression to Long COVID rather than to SARS-CoV-2 infection alone. In contrast, these comorbid conditions were rare among control patients without hEDS, irrespective of Long COVID status (**Figure 6b**). ME/CFS was the only condition observed in more than 20 control patients, affecting about 4% of controls, while all other single or combined comorbidity patterns were infrequent. Among control patients with Long COVID, 22.8% had a diagnosis of ME/CFS.

Together, these results indicate that Long COVID in hEDS is characterized by a high load of autonomic, immune, and fatigue-related comorbidities, with significant multimorbidity enrichment compared to the broad hEDS population, highlighting a role for multisystem vulnerability in post-viral sequelae.

### hEDS is associated with elevated risk factors for Long COVID and severe infection

To determine whether patients with hEDS are more susceptible to clinical conditions that increase the risk for severe COVID-19 outcomes and Long COVID, we compared ORs for various conditions between the hEDS cohort and matched controls using conditional logistic regression with Benjamini–Hochberg false discovery rate correction (**Figure 7a**). Patients with hEDS exhibited significantly increased odds for several diagnostic categories previously associated with worse outcomes after COVID-19 infection. All associations described below remain statistically significant after FDR correction (q < 0.05).

**Figure 7.**
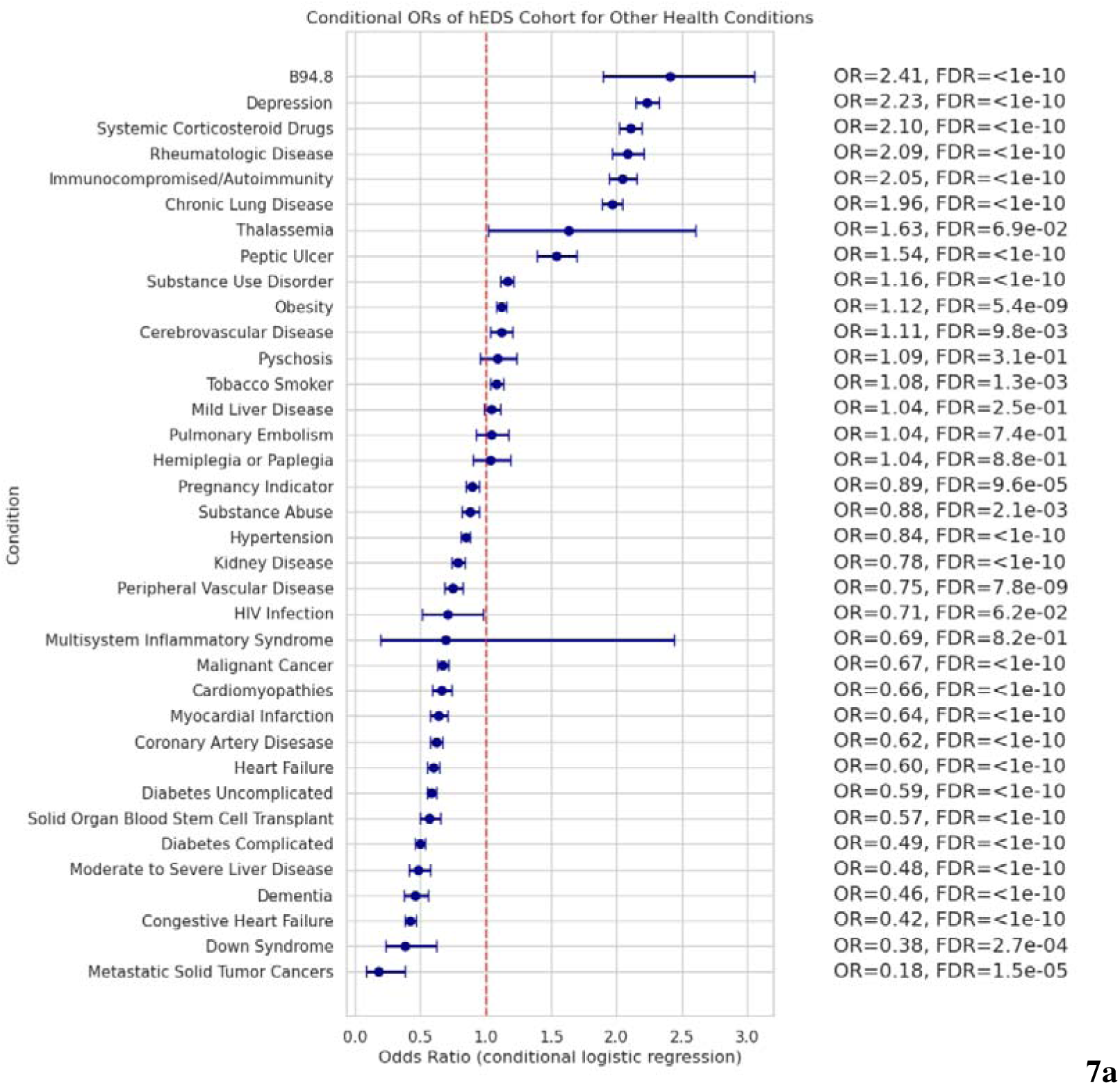
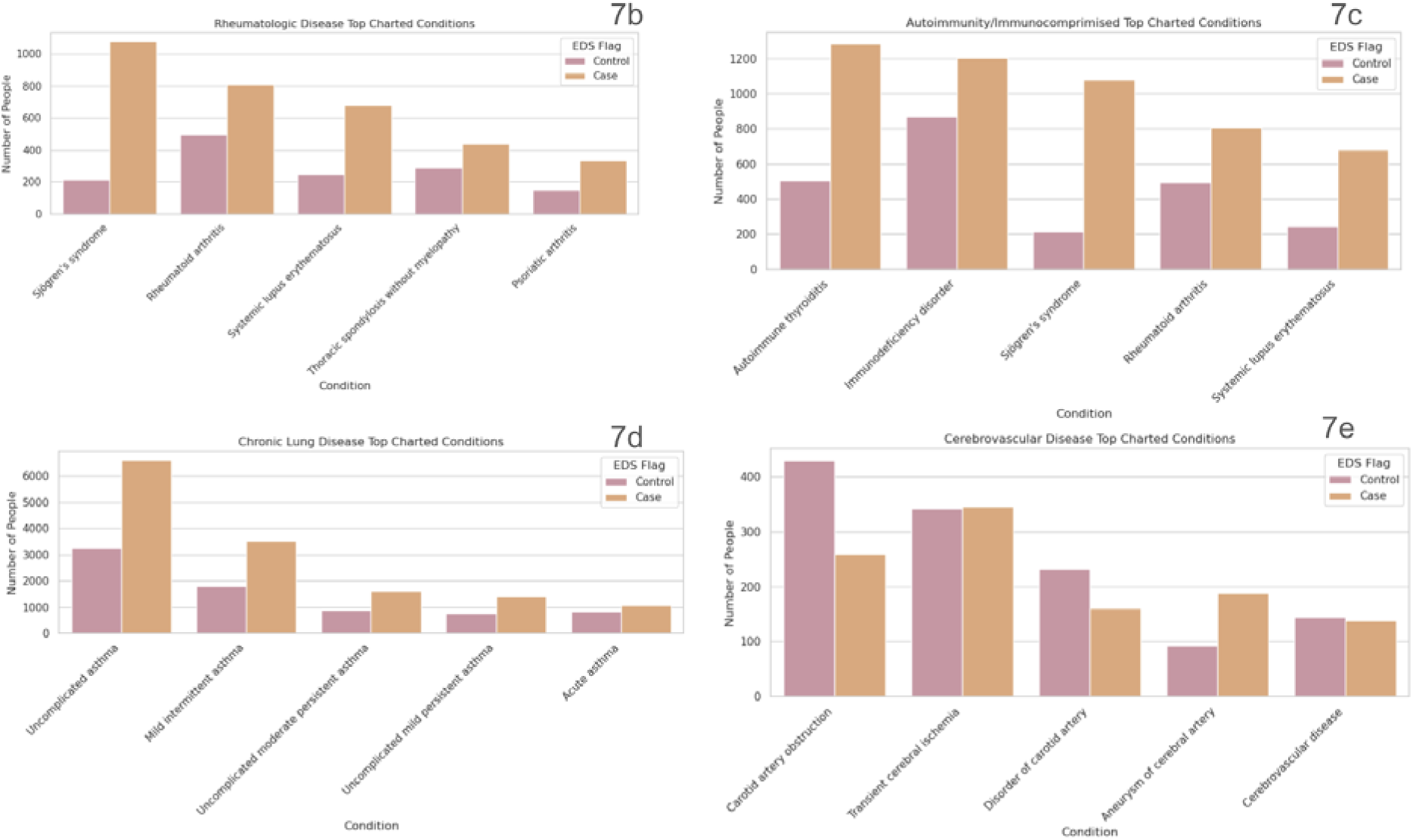
Patients with hEDS have increased odds related to COVID-19 categories. (7a) Odds ratios (ORs) with 95% confidence intervals (CIs) are shown for COVID-19–related diagnostic categories and known risk factors for severe infection in patients with hEDS compared to matched controls. Multiple testing was addressed using Benjamini–Hochberg false discovery rate correction; FDR-adjusted q values are shown in the figure. Patients with hEDS demonstrated elevated odds for several infectious and chronic conditions, as well as commonly co-occurring hEDS conditions. **(7b–e)** Raw counts of top contributing diagnoses within selected enriched categories are shown: **(7b)** rheumatologic disease, **(7c)** immunocompromised/autoimmunity, **(7d)** chronic lung disease, and **(7e)** cerebrovascular disease. All patient counts fewer than 20 are reported as “<20” to protect confidentiality. Additional contributing conditions are shown in **Supplementary Figure 4**.

Among the most striking associations were diagnoses categorized under sequelae of infectious disease, most commonly coded using B94.8, which was frequently used to document post-viral complications including early Long COVID, were more common in the hEDS group (OR = 2.41; 95% CI: 1.90-3.05). Patients with hEDS were also substantially more likely to have diagnoses of rheumatologic diseases (OR = 2.09; 95% CI: 1.97-2.21) and autoimmune disorders (OR = 2.05; 95% CI:1.94-2.15), both of which have been associated with increased risk of post-viral syndromes and pro-inflammatory responses to infection.

Chronic lung diseases were significantly elevated among patients with hEDS (OR = 1.96; 95% CI:1.88-2.04), as were cerebrovascular disease (OR = 1.11; 95% CI:1.03-1.20; q = 9.8 10^-3^) potentially reflecting underlying connective tissue fragility in vascular and pulmonary systems. Psychosocial and treatment-related risk factors also showed increased odds, including depression (OR = 2.23; 95% CI:2.15-2.32) and systemic corticosteroid use (OR = 2.10; 95% CI: 2.02-2.19). A smaller but not statistically significant elevation was observed for thalassemia (OR = 1.63; 95% CI: 1.02-2.61; q = 0.069). Finally, peptic ulcers were significantly elevated in patients with hEDS (OR = 1.54; 95% CI: 1.39-1.70). In contrast, several traditional risk factors for COVID-19 severity, such as diabetes, hypertension, liver disease, HIV, and cancer did not show significant enrichment among patients with hEDS.

To further characterize the source of elevated odds, we examined condition-level data to identify the specific diagnoses that most strongly contribute to each risk category (**Figures 7b–e**). The sequelae of infectious disease were primarily linked to ICD code B94.8, with 233 patients with hEDS having this code versus 97 controls. The rise in rheumatologic disease among patients with hEDS was largely driven by Sjögren’s syndrome (1,078 hEDS vs. 214 controls), rheumatoid arthritis (807 vs. 493), and systemic lupus erythematosus (680 vs. 246), along with contributions from thoracic spondylosis and psoriatic arthritis. In the autoimmunity category, autoimmune thyroiditis (1,284 vs. 505) and immunodeficiency disorders (1,204 vs. 870) were the most prevalent. The increased odds of chronic lung disease were largely due to asthma and chronic obstructive pulmonary disease (COPD). For cerebrovascular disease, carotid artery obstruction, and cerebral aneurysms were among the most common diagnoses.

These findings reveal that patients with hEDS carry a heightened burden of pre-existing conditions linked to worse viral outcomes and suggest a greater biological vulnerability to severe infection and persistent post-viral syndromes such as Long COVID. Additional condition-level details are available in **Supplementary Figure 4**.

## Discussion

In this study, we found that the prevalence of hEDS in the N3C cohort is approximately 1 in 800 individuals, which is higher than earlier estimates, though this may reflect both growing clinical awareness of hEDS and characteristics of the N3C cohort. To our knowledge, this represents the first large-scale, multi-site characterization of hEDS in the United States using real-world EHR data. Clinical validation through chart review was not possible, limiting interpretation; false positives or misclassifications could occur due to rule-out diagnoses being coded as confirmed, post-2017 criteria expansion, or diagnostic substitution during the Long Covid era. Future research should include chart-based validation to confirm these findings. Given the complexity and under-recognition of hEDS, the true prevalence is likely even higher, reinforcing the need for increased clinical awareness, diagnostic support tools, and further prevalence studies in other populations. Further characterization of the hEDS cohort revealed a common demographic profile of White, non-Hispanic females between the ages of 25–45, consistent with prior studies[24]. While this may reflect actual patterns of disease distribution, it might also indicate disparities in access to specialty care and diagnosis. More research is needed to understand demographic differences in symptom presentation, care-seeking behavior, and clinical recognition, as well as to identify and support underserved populations who may be at risk for missing diagnoses.

In examining the relationship between hEDS, COVID-19, and Long COVID, we found that while rates of confirmed COVID-19 infection were similar between patients with hEDS and matched controls, patients with hEDS exhibited significantly higher odds of Long COVID diagnoses and Long COVID-related clinic visits. This could reflect symptom exacerbation, underlying pathophysiological overlap, or increased healthcare engagement and diagnostic attention following infection. We also observed elevated use of ICD-10 code B94.8, “sequelae of infectious disease” in patients with hEDS, which was widely used as a proxy for Long COVID prior to the release of a dedicated Long COVID code (U09.9) in late 2021. However, this code was also often applied to pre-existing post-viral symptoms, highlighting limitations of code-based inference. Further follow-up of the timing of this diagnostic code is necessary to reach a definitive conclusion. Overall, these findings suggest that patients with hEDS are particularly vulnerable to developing Long COVID after a COVID-19 infection, which we speculate could be due to symptom exacerbation or underlying pathophysiological overlap, though alternative explanations cannot be excluded.

We also found that patients with hEDS were more likely to receive outpatient treatments for COVID-19, such as Paxlovid, but not Remdesivir, which is administered intravenously and typically used in inpatient settings. This pattern may reflect a higher likelihood of moderate complications requiring attention, differences in provider prescribing practices, or the relative accessibility of oral versus intravenous treatments. Additionally, Paxlovid became available in late 2021, as awareness of Long COVID risk increased, so clinicians might preferentially prescribe Paxlovid to patients with known chronic conditions such as hEDS. To fully understand these results, further investigation into hospitalization rates or ICU admissions would be necessary.

Increases in hEDS diagnoses during the COVID-19 pandemic raise concerns about classical ascertainment bias caused by increased healthcare utilization. Although raw diagnosis counts showed different patterns over time across COVID-19 exposure groups, encounter-adjusted hEDS diagnosis rates remained stable over time and did not vary by COVID-19 or Long COVID status. Under a typical ascertainment bias model, increased healthcare contact, especially after COVID-19 or Long COVID, would lead to higher diagnosis rates per encounter. The lack of such a rise suggests that the changes seen in raw counts are due to shifts in healthcare utilization and diagnostic timing rather than an actual increase in diagnostic yield. While some bias related to encounter type or referral patterns may still exist, these results argue against classical utilization-driven ascertainment bias being the main reason for the observed temporal trends in hEDS diagnosis. Nevertheless, residual bias related to encounter type, referral patterns, or coding practices cannot be excluded.

Comorbid conditions commonly observed in hEDS, including MCAS, POTS, and ME/CFS, were significantly more prevalent in patients with hEDS with Long COVID than in those without. While these comorbidities were not more common in patients with hEDS who contracted COVID-19 compared to those who did not, some COVID-19 infections might have gone unrecorded due to home testing or incomplete charting. Notably, cumulative incidence analyses revealed that hEDS diagnoses were more likely to occur after a COVID-19 or Long COVID diagnosis, particularly among patients with MCAS and POTS. This suggests that viral infections may serve as unmasking events, prompting more thorough clinical investigation and eventual recognition of hEDS. For example, post-viral worsening of autonomic symptoms or immune dysregulation may lead clinicians to consider diagnoses such as POTS or MCAS, which can then reveal the underlying connective tissue disorder. However, alternative explanations, including increased healthcare contact and diagnostic attention, remain plausible.

Nearly all patients with hEDS with MCAS who were later diagnosed with hEDS received that diagnosis after a COVID-19 infection. These findings support recent hypotheses that hypermobile connective tissue disorders may predispose individuals to immune dysregulation through mast cell activation, potentially contributing to both hEDS symptoms and Long COVID risk[36]. A similar pattern was observed for patients with POTS, suggesting that post-viral autonomic dysfunction may also act as a trigger for hEDS diagnosis. Conversely, the cumulative incidence of COVID-19 or Long COVID after an hEDS diagnosis was lower, suggesting that while hEDS may not increase susceptibility to infection, it probably raises the risk of developing prolonged post-viral complications.

In addition to vulnerabilities specific to Long COVID, we observed significantly higher odds for a variety of chronic conditions in the hEDS cohort, including rheumatologic disease, autoimmune disease, chronic lung disease, cerebrovascular disease, depression, systemic corticosteroid use, thalassemia, and peripheral vascular disease. Some of these, such as autoimmune conditions and depression, are well-documented hEDS comorbidities. Others, like thalassemia and peptic ulcers, are less commonly reported, and may reflect true comorbidities, coding artifacts, or misclassifications; these findings are exploratory. The observed elevation in peptic ulcers, which are not typically associated with hEDS, warrants further study. The increased use of systemic corticosteroids is also notable because they may negatively affect the integrity of the connective tissue; such treatments may precede hEDS diagnosis or exacerbate undiagnosed symptoms, leading to a diagnostic investigation, though causal interpretation is speculative.

## Limitations

Misclassification is an inherent limitation of registry-based analyses of hEDS. Dysautonomia, POTS, MCAS, and nonspecific EDS codes may be inconsistently or incorrectly recorded as hEDS, especially during periods of healthcare disruption such as the COVID-19 pandemic. Additionally, differences in coding practices between institutions and incomplete longitudinal documentation can lower diagnostic accuracy, even when using established code-based definitions.

COVID-19 infection rates declined over time, consistent with population-level trends related to vaccination, prior immunity, and changes in testing practices. Identification of Long COVID is inherently dependent on prior SARS-CoV-2 infection; however, COVID-19 infections may be underascertained or inconsistently documented in EHR data, particularly for early-pandemic or externally tested cases. This likely results in misclassification and underestimation of Long COVID incidence.

Certain rare diagnoses, such as vertebral tuberculosis, were identified in the dataset but are not shown in figures or analyses. Upon review, these codes originated from multiple clinical sites with no consistent pattern, and the observed associations were implausibly high considering how rare vertebral tuberculosis is in the U.S. We believe these entries are likely coding errors or misclassifications (e.g., lower back pain coded as vertebral TB). Because we cannot interpret these data reliably, they were excluded from the figures and analyses, which highlights a limitation of relying solely on diagnosis codes.

The comparable condition analyses are limited, as the selection of alternative disorders is restricted by both sample size and co-occurrence with hEDS. Future work is needed to validate findings using larger or more diverse comparator groups.

Cox and time-to-event analyses also have important limitations. Comorbid conditions were defined based on documentation at any time during follow-up and therefore may occur after the COVID-19 index date or after hEDS diagnosis; as such, these variables should be interpreted as markers of phenotypic clustering and healthcare interaction rather than temporally ordered risk factors. Covariates were modeled as fixed indicators rather than fully time-varying exposures. Administrative censoring was based on the latest available data cutoff and may vary across individuals. Diagnostic timing reflects documentation rather than true disease onset, and residual confounding by healthcare utilization is likely.

Additional limitations include the inability to review charts and validate diagnoses clinically, which prevents confirmation of true hEDS status. Date-shifting in N3C can introduce potential measurement error for short-term temporal analyses, although person-level ordering of events preserves the relative timing of COVID-19 exposure and hEDS diagnosis. Finally, rare conditions and post-infection outcomes should be interpreted carefully, as higher odds may reflect coding artifacts, healthcare utilization patterns, or residual confounding rather than actual biological susceptibility.

## Conclusions

This study presents the largest real-world characterization of hEDS in the United States to date, using a harmonized, multi-institutional electronic health record dataset. We estimate the prevalence of hEDS to be approximately 1 in 800 individuals, indicating the condition is far more common than previously thought and remains significantly underdiagnosed.

Patients with hEDS demonstrated significantly elevated rates of comorbid conditions, particularly MCAS, POTS, and ME/CFS, which were also associated with an increased likelihood of developing Long COVID. ORs for other rheumatologic and neurovascular conditions were also elevated, underscoring the systemic complexity of hEDS and its potential interactions with immune and autonomic pathways.

This data supports the hypothesis that stress from immune activation after viral infections, such as Long COVID in this study, may cause patients with an underlying connective tissue disorder to receive a diagnosis after symptom worsening following viral infection. Cumulative incidence analyses revealed that many hEDS diagnoses happened after COVID-19 or Long COVID diagnoses, suggesting that viral illness may unmask previously unrecognized hEDS symptoms. These findings highlight the need for integrated, multidisciplinary care models for individuals with hEDS, particularly in the context of post-viral syndromes like Long COVID. Increased clinical awareness, better diagnostic strategies, and more research into the pathophysiological overlap between hEDS and viral infections are essential to ensure timely diagnosis, management, and support for this vulnerable patient population.

## Data Availability

This study used de-identified Level 2 electronic health record data from the National Clinical Cohort Collaborative (N3C) COVID-19 Enclave, which includes date-shifted patient data for privacy protection. The data used are not publicly available due to institutional, federal, and patient privacy regulations, but they are accessible to qualified researchers through an application process. Researchers may apply for data access by submitting a Data Use Request (DUR) through the NCATS N3C Data Enclave and completing necessary data use and security training requirements. All analyses in this study were conducted under approved Data Use Request RP-582482. The authors did not have any special privileges in accessing the data. Access to the same dataset used in this manuscript is available to other researchers following the same application procedures.

https://covid.cd2h.org/enclave

## Data and Code Availability Statement

All analysis code used in this study is publicly available at GitHub (https://github.com/megankraus/hEDS_covid) and archived on Zenodo (https://doi.org/10.5281/zenodo.18508715).

Analyses were conducted within the National COVID Cohort Collaborative (N3C) Enclave using de-identified electronic health record data mapped to the OMOP Common Data Model. Due to N3C governance restrictions, raw patient-level data cannot be shared. Researchers with approved N3C access can reproduce all analyses using the provided code.

## Acknowledgments, Competing Interests, Funding and all other required statements

### Authorship

Authorship was determined using ICMJE recommendations.

### N3C Attribution

The analyses described in this publication were conducted with data or tools accessed through the N3C Data Enclave. This research was possible because of the patients whose information is included within the data and the organizations (https://ncats.nih.gov/n3c/resources/data-contribution/data-transfer-agreement-signatories) and scientists who have contributed to the on-going development of this community resource [https://doi.org/10.1093/jamia/ocaa196].

### Disclaimer

The N3C Publication committee confirmed that this manuscript msid:2549.961 is in accordance with N3C data use and attribution policies; however, this content is solely the responsibility of the authors and does not represent the official views of the National Institutes of Health or the N3C program.

### IRB

The N3C data transfer to NCATS is performed under a Johns Hopkins University Reliance Protocol # IRB00249128 or individual site agreements with NIH. The N3C Data Enclave is managed under the authority of the NIH; information can be found at https://ncats.nih.gov/n3c/resources.

## Acknowledgements

We gratefully acknowledge Johanna J. Loomba and Shawn T. O’Neil for their contributions for developing Logic Liaison templates and training materials.

## Data Partners with Released Data

The following institutions whose data is released or pending:

Available: Advocate Health Care Network — UL1TR002389: The Institute for Translational Medicine (ITM) • Aurora Health Care Inc — UL1TR002373: Wisconsin Network For Health Research • Boston University Medical Campus — UL1TR001430: Boston University Clinical and Translational Science Institute • Brown University — U54GM115677: Advance Clinical Translational Research (Advance-CTR) • Carilion Clinic — UL1TR003015: iTHRIV Integrated Translational health Research Institute of Virginia • Case Western Reserve University — UL1TR002548: The Clinical & Translational Science Collaborative of Cleveland (CTSC) • Charleston Area Medical Center — U54GM104942: West Virginia Clinical and Translational Science Institute (WVCTSI) • Children’s Hospital Colorado — UL1TR002535: Colorado Clinical and Translational Sciences Institute • Columbia University Irving Medical Center — UL1TR001873: Irving Institute for Clinical and Translational Research • Dartmouth College — None (Voluntary) Duke University — UL1TR002553: Duke Clinical and Translational Science Institute • George Washington Children’s Research Institute — UL1TR001876: Clinical and Translational Science Institute at Children’s National (CTSA-CN) • George Washington University — UL1TR001876: Clinical and Translational Science Institute at Children’s National (CTSA-CN) • Harvard Medical School — UL1TR002541: Harvard Catalyst • Indiana University School of Medicine — UL1TR002529: Indiana Clinical and Translational Science Institute • Johns Hopkins University — UL1TR003098: Johns Hopkins Institute for Clinical and Translational Research • Louisiana Public Health Institute — None (Voluntary) • Loyola Medicine — Loyola University Medical Center • Loyola University Medical Center — UL1TR002389: The Institute for Translational Medicine (ITM) • Maine Medical Center — U54GM115516: Northern New England Clinical & Translational Research (NNE-CTR) Network • Mary Hitchcock Memorial Hospital & Dartmouth Hitchcock Clinic — None (Voluntary) • Massachusetts General Brigham — UL1TR002541: Harvard Catalyst • Mayo Clinic Rochester — UL1TR002377: Mayo Clinic Center for Clinical and Translational Science (CCaTS) • Medical University of South Carolina — UL1TR001450: South Carolina Clinical & Translational Research Institute (SCTR) • MITRE Corporation — None (Voluntary) • Montefiore Medical Center — UL1TR002556: Institute for Clinical and Translational Research at Einstein and Montefiore • Nemours — U54GM104941: Delaware CTR ACCEL Program • NorthShore University HealthSystem — UL1TR002389: The Institute for Translational Medicine (ITM) • Northwestern University at Chicago — UL1TR001422: Northwestern University Clinical and Translational Science Institute (NUCATS) • OCHIN — INV-018455: Bill and Melinda Gates Foundation grant to Sage Bionetworks • Oregon Health & Science University — UL1TR002369: Oregon Clinical and Translational Research Institute • Penn State Health Milton S. Hershey Medical Center — UL1TR002014: Penn State Clinical and Translational Science Institute • Rush University Medical Center — UL1TR002389: The Institute for Translational Medicine (ITM) • Rutgers, The State University of New Jersey — UL1TR003017: New Jersey Alliance for Clinical and Translational Science • Stony Brook University — U24TR002306 • The Alliance at the University of Puerto Rico, Medical Sciences Campus — U54GM133807: Hispanic Alliance for Clinical and Translational Research (The Alliance) • The Ohio State University — UL1TR002733: Center for Clinical and Translational Science • The State University of New York at Buffalo — UL1TR001412: Clinical and Translational Science Institute • The University of Chicago — UL1TR002389: The Institute for Translational Medicine (ITM) • The University of Iowa — UL1TR002537: Institute for Clinical and Translational Science • The University of Miami Leonard M. Miller School of Medicine — UL1TR002736: University of Miami Clinical and Translational Science Institute • The University of Michigan at Ann Arbor — UL1TR002240: Michigan Institute for Clinical and Health Research • The University of Texas Health Science Center at Houston — UL1TR003167: Center for Clinical and Translational Sciences (CCTS) • The University of Texas Medical Branch at Galveston — UL1TR001439: The Institute for Translational Sciences • The University of Utah — UL1TR002538: Uhealth Center for Clinical and Translational Science • Tufts Medical Center — UL1TR002544: Tufts Clinical and Translational Science Institute • Tulane University — UL1TR003096: Center for Clinical and Translational Science • The Queens Medical Center — None (Voluntary) • University Medical Center New Orleans — U54GM104940: Louisiana Clinical and Translational Science (LA CaTS) Center • University of Alabama at Birmingham — UL1TR003096: Center for Clinical and Translational Science • University of Arkansas for Medical Sciences — UL1TR003107: UAMS Translational Research Institute • University of Cincinnati — UL1TR001425: Center for Clinical and Translational Science and Training • University of Colorado Denver, Anschutz Medical Campus — UL1TR002535: Colorado Clinical and Translational Sciences Institute • University of Illinois at Chicago — UL1TR002003: UIC Center for Clinical and Translational Science • University of Kansas Medical Center — UL1TR002366: Frontiers: University of Kansas Clinical and Translational Science Institute • University of Kentucky — UL1TR001998: UK Center for Clinical and Translational Science • University of Massachusetts Medical School Worcester — UL1TR001453: The UMass Center for Clinical and Translational Science (UMCCTS) • University Medical Center of Southern Nevada — None (voluntary) • University of Minnesota — UL1TR002494: Clinical and Translational Science Institute • University of Mississippi Medical Center — U54GM115428: Mississippi Center for Clinical and Translational Research (CCTR) • University of Nebraska Medical Center — U54GM115458: Great Plains IDeA-Clinical & Translational Research • University of North Carolina at Chapel Hill — UL1TR002489: North Carolina Translational and Clinical Science Institute • University of Oklahoma Health Sciences Center — U54GM104938: Oklahoma Clinical and Translational Science Institute (OCTSI) • University of Pittsburgh — UL1TR001857: The Clinical and Translational Science Institute (CTSI) • University of Pennsylvania — UL1TR001878: Institute for Translational Medicine and Therapeutics • University of Rochester — UL1TR002001: UR Clinical & Translational Science Institute • University of Southern California — UL1TR001855: The Southern California Clinical and Translational Science Institute (SC CTSI) • University of Vermont — U54GM115516: Northern New England Clinical & Translational Research (NNE-CTR) Network • University of Virginia — UL1TR003015: iTHRIV Integrated Translational health Research Institute of Virginia • University of Washington — UL1TR002319: Institute of Translational Health Sciences • University of Wisconsin-Madison — UL1TR002373: UW Institute for Clinical and Translational Research • Vanderbilt University Medical Center — UL1TR002243: Vanderbilt Institute for Clinical and Translational Research • Virginia Commonwealth University — UL1TR002649: C. Kenneth and Dianne Wright Center for Clinical and Translational Research • Wake Forest University Health Sciences — UL1TR001420: Wake Forest Clinical and Translational Science Institute • Washington University in St. Louis — UL1TR002345: Institute of Clinical and Translational Sciences • Weill Medical College of Cornell University — UL1TR002384: Weill Cornell Medicine Clinical and Translational Science Center • West Virginia University — U54GM104942: West Virginia Clinical and Translational Science Institute (WVCTSI)□ Submitted: Icahn School of Medicine at Mount Sinai — UL1TR001433: ConduITS Institute for Translational Sciences • The University of Texas Health Science Center at Tyler — UL1TR003167: Center for Clinical and Translational Sciences (CCTS) • University of California, Davis — UL1TR001860: UCDavis Health Clinical and Translational Science Center • University of California, Irvine — UL1TR001414: The UC Irvine Institute for Clinical and Translational Science (ICTS) • University of California, Los Angeles — UL1TR001881: UCLA Clinical Translational Science Institute • University of California, San Diego — UL1TR001442: Altman Clinical and Translational Research Institute • University of California, San Francisco — UL1TR001872: UCSF Clinical and Translational Science Institute□ NYU Langone Health Clinical Science Core, Data Resource Core, and PASC Biorepository Core — OTA-21-015A: Post-Acute Sequelae of SARS-CoV-2 Infection Initiative (RECOVER)□ Pending: Arkansas Children’s Hospital — UL1TR003107: UAMS Translational Research Institute • Baylor College of Medicine — None (Voluntary) • Children’s Hospital of Philadelphia — UL1TR001878: Institute for Translational Medicine and Therapeutics • Cincinnati Children’s Hospital Medical Center — UL1TR001425: Center for Clinical and Translational Science and Training • Emory University — UL1TR002378: Georgia Clinical and Translational Science Alliance • HonorHealth — None (Voluntary) • Loyola University Chicago — UL1TR002389: The Institute for Translational Medicine (ITM) • Medical College of Wisconsin — UL1TR001436: Clinical and Translational Science Institute of Southeast Wisconsin • MedStar Health Research Institute — None (Voluntary) • Georgetown University — UL1TR001409: The Georgetown-Howard Universities Center for Clinical and Translational Science (GHUCCTS) • MetroHealth — None (Voluntary) • Montana State University — U54GM115371: American Indian/Alaska Native CTR • NYU Langone Medical Center — UL1TR001445: Langone Health’s Clinical and Translational Science Institute • Ochsner Medical Center — U54GM104940: Louisiana Clinical and Translational Science (LA CaTS) Center • Regenstrief Institute — UL1TR002529: Indiana Clinical and Translational Science Institute • Sanford Research — None (Voluntary) • Stanford University — UL1TR003142: Spectrum: The Stanford Center for Clinical and Translational Research and Education • The Rockefeller University — UL1TR001866: Center for Clinical and Translational Science • The Scripps Research Institute — UL1TR002550: Scripps Research Translational Institute • University of Florida — UL1TR001427: UF Clinical and Translational Science Institute • University of New Mexico Health Sciences Center — UL1TR001449: University of New Mexico Clinical and Translational Science Center • University of Texas Health Science Center at San Antonio — UL1TR002645: Institute for Integration of Medicine and Science • Yale New Haven Hospital — UL1TR001863: Yale Center for Clinical Investigation

## Code Availability and Reproducibility

The full analysis pipeline, including Spark SQL, R, and Python nodes executed in the N3C Enclave, is publicly available at GitHub and archived on Zenodo (DOI: 10.5281/zenodo.18508715). The repository documents execution order, cohort definitions, and N3C-specific implementation details. All analyses are deterministic given the same N3C data release and approved access.

## Author Contributions

Conceptualization MLP, GB, MHA

Data curation: MLP, BJL

Formal analysis: MLP

Funding acquisition: GB, MHA

Investigation: MLP, ERE

Methodology: MLP, BJL, MHA

Project administration: GB, MHA

Supervision: GB, ERE, MHA

Visualization: MLP

Writing – original draft: MLP

Writing – review & editing: All authors

## Supplementary

**Supplementary Table 1. OMOP concept IDs used for cohort definitions.**

Listed are condition-level OMOP concept identifiers used to define hEDS, other Ehlers-Danlos syndrome subtypes, MCAS, POTS, ME/CFS, COVID-19, and post-acute COVID-19 (Long COVID).

**Supplementary Table 2. Logic Liaison conditions.**

Listed are Logic Liaison–derived condition, diagnosis, medication, and laboratory codeset identifiers used to define comorbidities, COVID-19–related outcomes, treatments, and testing status in the study.

**Supplementary Table 3: Covariates of Cases and Controls**

Showing raw counts for categorical variables between cases (hEDS) and controls. Continuous variables are averaged, with standard deviation shown. All counts <20 are shown as such for patient protection. Data partner IDs are obfuscated for clinical site protection.

**Supplementary Table 4: Odds ratio values and statistics**

This table presents the odds ratios (ORs) and 95% confidence intervals (CIs) for each condition comparing patients with hEDS to matched controls. FDR-adjusted q-values are also provided to account for multiple testing across all conditions. ORs with q < 0.05 are considered statistically significant after false discovery rate correction. Analyses were performed using conditional logistic regression accounting for matched pairs and healthcare utilization.

**Supplementary Table 5: Prevalence Rates of Comorbidities Segmented by Long COVID Flag.** This table shows the prevalence of individual comorbidities and their combinations among patients stratified by hEDS diagnosis (EDS flag) and Long COVID status (Long COVID flag). Prevalence is reported as the proportion of patients with each condition within the respective subgroup. Combinations of comorbidities are included to highlight patterns of co-occurrence. These data provide context for phenotypic clustering and potential interactions between hEDS and Long COVID.

**Supplemental Table 6: Cumulative incidence of hEDS diagnoses by comorbidity group and index condition.** This table presents cumulative incidence (%) of hEDS diagnoses at 180, 365, and 730 days for patients stratified by comorbidity group (POTS, MCAS, ME/CFS, or none) and index condition (COVID-19 or Long COVID). Log-rank p-values are shown for pairwise comparisons between each comorbidity group and reference groups. A dash (“–”) indicates comparison with the same group. These data provide insight into the temporal patterns of hEDS diagnosis across comorbidities and infection status.

**Supplemental Figure 1. Prevalence of comorbid conditions in COVID-19 patients by hEDS status.** Bar heights represent the prevalence of commonly reported comorbid conditions among patients with hEDS and COVID-19 (n = 25,152) versus matched controls with COVID-19 (n = 25,152). Prevalence for combinations of conditions is also shown. To protect patient confidentiality, counts fewer than 20 are reported as <20 or left blank. Overall, patients with hEDS exhibit higher prevalence of these comorbidities, suggesting that pre-existing conditions may contribute to risk of complications following COVID-19.

**Supplemental Figure 2. Interrupted time series of monthly hEDS diagnosis counts (2018–2026).** Monthly counts of hEDS diagnoses are shown from January 2018 through 2026. The vertical dashed line indicates the start of the COVID-19 pandemic (January 2020), and segmented regression lines illustrate the trend before and after this interruption. This analysis allows visualization of temporal changes in hEDS diagnoses over time and the potential impact of the pandemic on diagnostic patterns.

**Supplemental Figure 3: Cumulative incidence plots show patients with hEDS have higher likelihood of receiving a diagnosis after an acute post viral infection with complications when comorbidities are present.** Cumulative incidence plots demonstrate that patients with hEDS have a higher likelihood of receiving a diagnosis following acute post-viral infection and experience greater risk of complications when comorbid conditions are present. Panels **3a and c** show the cumulative incidence of receiving an hEDS diagnosis after COVID-19 infection (**3a)** or Long COVID (**3c)**. Panels **3b and d** assess the cumulative incidence of developing either a COVID-19 infection (**3b)** or Long COVID (**3d)** after receiving an hEDS diagnosis. Time is measured in days from the index event (e.g., COVID-19 diagnosis or hEDS diagnosis), and cumulative incidence was estimated using the Kaplan-Meier method. Shaded regions represent 95% confidence intervals.

**Supplemental Figure 4. Additional Top Charted Conditions.** Raw counts of top contributing diagnoses of remaining categories are shown: **(4a)** pulmonary embolism, **(4b)** peripheral vascular disease **(4c)** systemic corticosteroids **(4d)** depression, **(4e)** peptic ulcer, and **(4f)** B94.8. All patient counts fewer than 20 are reported as “<20” to protect confidentiality.

## Citations

1. Miklovic T, Sieg. VC. Ehlers-Danlos Syndrome. In: StatPearls [Internet]. 29 May 2023 [cited 3 Jun 2025]. Available: https://www.ncbi.nlm.nih.gov/books/NBK549814/

2. Malfait F, Francomano C, Byers P, Belmont J, Berglund B, Black J, et al. The 2017 international classification of the Ehlers-Danlos syndromes. Am J Med Genet C Semin Med Genet. 2017;175: 8–26.

3. Blackburn PR, Xu Z, Tumelty KE, Zhao RW, Monis WJ, Harris KG, et al. Bi-allelic alterations in AEBP1 lead to defective collagen assembly and connective tissue structure resulting in a variant of Ehlers-Danlos syndrome. Am J Hum Genet. 2018;102: 696–705.

4. Hypermobile Ehlers-Danlos syndrome (a.k.. Ehlers-Danlos syndrome Type III Ehlers-Danlos syndrome hypermobility type): clinical description natural history.

5. Petrucci T, Barclay SJ, Gensemer C, Morningstar J, Daylor V, Byerly K, et al. Phenotypic clusters and multimorbidity in hypermobile Ehlers-Danlos syndrome. Mayo Clin Proc Innov Qual Outcomes. 2024;8: 253–262.

6. Fikree A, Chelimsky G, Collins H, Kovacic K, Aziz Q. Gastrointestinal involvement in the Ehlers-Danlos syndromes. Am J Med Genet C Semin Med Genet. 2017;175: 181–187.

7. Gullapalli PA, Javed S. Multidisciplinary chronic pain management strategies in patients with Ehlers-Danlos syndromes. Pain Manag. 2023;13: 5–14.

8. Chopra P, Tinkle B, Hamonet C, Brock I, Gompel A, Bulbena A, et al. Pain management in the Ehlers-Danlos syndromes. Am J Med Genet C Semin Med Genet. 2017;175: 212–219.

9. Hakim A, O’Callaghan C, De Wandele I, Stiles L, Pocinki A, Rowe P. Cardiovascular autonomic dysfunction in Ehlers-Danlos syndrome-Hypermobile type. Am J Med Genet C Semin Med Genet. 2017;175: 168–174.

10. Mathias CJ, Owens A, Iodice V, Hakim A. Dysautonomia in the Ehlers-Danlos syndromes and hypermobility spectrum disorders-With a focus on the postural tachycardia syndrome. Am J Med Genet C Semin Med Genet. 2021;187: 510–519.

11. Darakjian AA, Bhutani M, Fairweather D, Kocsis SC, Fliess JJ, Khatib S, et al. Similarities and differences in self-reported symptoms and comorbidities between hypermobile Ehlers-Danlos syndrome and hypermobility spectrum disorders. Rheumatol Adv Pract. 2024;8: rkae134.

12. People O. HEDS. In: The Ehlers Danlos Society [Internet]. 2 Jul 2024 [cited 3 Jun 2025]. Available: https://www.ehlers-danlos.com/heds/

13. People O. What is HSD? In: The Ehlers Danlos Society [Internet]. 15 Mar 2017 [cited 18 Jun 2025]. Available: https://www.ehlers-danlos.com/what-is-hsd/

14. Thwaites PA, Gibson PR, Burgell RE. Hypermobile Ehlers-Danlos syndrome and disorders of the gastrointestinal tract: What the gastroenterologist needs to know. J Gastroenterol Hepatol. 2022;37: 1693–1709.

15. Ishiguro H, Yagasaki H, Horiuchi Y. Ehlers-Danlos syndrome in the field of psychiatry: A review. Front Psychiatry. 2021;12: 803898.

16. Seneviratne SL, Maitland A, Afrin L. Mast cell disorders in Ehlers-Danlos syndrome. Am J Med Genet C Semin Med Genet. 2017;175: 226–236.

17. Farley M, Estrada-Mendizabal RJ, Gansert EA, Voelker D, Marks LA, Gonzalez-Estrada A. Prevalence of mast cell activation disorders and hereditary alpha tryptasemia among patients with postural orthostatic tachycardia syndrome and Ehlers-Danlos syndrome: A systematic review. Ann Allergy Asthma Immunol. 2025. doi:10.1016/j.anai.2025.03.022

18. Gulen T. Using the right criteria for MCAS. Curr Allergy Asthma Rep. 2024;24: 39–51.

19. Bonamichi-Santos R, Yoshimi-Kanamori K, Giavina-Bianchi P, Aun MV. Association of postural tachycardia syndrome and Ehlers-Danlos syndrome with mast cell activation disorders. Immunol Allergy Clin North Am. 2018;38: 497–504.

20. Anderson LK, Lane KR. Clinical trajectory of hypermobile Ehlers-Danlos syndrome/hypermobility spectrum disorders in older adults. J Am Assoc Nurse Pract. 2023;35: 605–612.

21. Re-Writing Natural History Pain Related Symptoms Joint Hypermobility Syndrome/ Ehlers-Danlos Syndrome, Hypermobility Type.

22. Schubart JR, Mills SE, Schaefer EW, Bascom R, Francomano CA. Longitudinal analysis of symptoms in the Ehlers-Danlos syndromes. Am J Med Genet A. 2022;188: 1204–1213.

23. Hugon-Rodin J, Lebègue G, Becourt S, Hamonet C, Gompel A. Gynecologic symptoms and the influence on reproductive life in 386 women with hypermobility type ehlers-danlos syndrome: a cohort study. Orphanet J Rare Dis. 2016;11: 124.

24. Demmler JC, Atkinson MD, Reinhold EJ, Choy E, Lyons RA, Brophy ST. Diagnosed prevalence of Ehlers-Danlos syndrome and hypermobility spectrum disorder in Wales, UK: a national electronic cohort study and case-control comparison. BMJ Open. 2019;9: e031365.

25. Wang Y-T, Jahani S, Morel-Swols D, Kapely A, Rosen A, Forghani I. Patient experiences of receiving a diagnosis of hypermobile Ehlers-Danlos syndrome. Am J Med Genet A. 2024;194: e63613.

26. Schubart JR, Schaefer EW, Knight DRT, Mills SE, Francomano CA. Estimates of the excess cost burden of Ehlers-Danlos syndromes: a United States MarketScan® claims database analysis. Front Public Health. 2024;12: 1365712.

27. Ganesh R, Munipalli B. Long COVID and hypermobility spectrum disorders have shared pathophysiology. Front Neurol. 2024;15: 1455498.

28. Eccles JA, Cadar D, Quadt L, Hakim AJ, Gall N, Covid Symptom Survey Biobank Consortium, et al. Is joint hypermobility linked to self-reported non-recovery from COVID-19? Case-control evidence from the British COVID Symptom Study Biobank. BMJ Public Health. 2024;2: e000478.

29. Grach SL, Dudenkov DV, Pollack B, Fairweather D, Aakre CA, Munipalli B, et al. Overlapping conditions in Long COVID at a multisite academic center. Front Neurol. 2024;15: 1482917.

30. Logarbo BP, Yang M, Longo MT, Kingry C, Courseault J. Long COVID and the diagnosis of underlying hypermobile Ehlers-Danlos syndrome and hypermobility spectrum disorders. PM R. 2024;16: 935–937.

31. agz5de. National-clinical-cohort-collaborative/logic-liaison-confirmed-COVID-positive-template: Template enhancements through may 2025. Zenodo; 2025. doi:10.5281/ZENODO.15556955

32. Ho DE, Imai K, King G, Stuart EA. MatchIt: Nonparametric Preprocessing for Parametric Causal Inference. J Stat Softw. 2011;42. doi:10.18637/jss.v042.i08

33. Leese P, Anand A, Girvin A, Manna A, Patel S, Yoo YJ, et al. Clinical encounter heterogeneity and methods for resolving in networked EHR data: a study from N3C and RECOVER programs. J Am Med Inform Assoc. 2023;30: 1125–1136.

34. Davidson-Pilon C. lifelines: survival analysis in Python. J Open Source Softw. 2019;4: 1317.

35. Hakim A. Hypermobile Ehlers-Danlos Syndrome. In: Adam MP, Bick S, Mirzaa GM, Pagon RA, Wallace SE, Amemiya A, editors. GeneReviews. Seattle (WA): University of Washington, Seattle; 2004.

36. Monaco A, Choi D, Uzun S, Maitland A, Riley B. Association of mast-cell-related conditions with hypermobile syndromes: a review of the literature. Immunol Res. 2022;70: 419–431.

