## Supplemental for "Understanding Comorbidities in Hypermobile Ehlers-Danlos Syndrome: Could a Viral Infection Unmask the Disorder?"

**Supplementary**

**Supplementary Table 1: Concept IDs for Cohort Criteria**

| Label | Type of Concept ID | OMOP ID |
| --- | --- | --- |
| Hypermobile Ehlers-Danlos Syndrome | Condition | 4148925 |
| General Ehlers-Danlos Syndrome | Condition | 79145 |
| Vascular Ehlers-Danlos Syndrome | Condition | 4062070 |
| Classical Ehlers-Danlos Syndrome | Condition | 37395762 |
| MCAS | Condition | 37309640 |
| POTS | Condition | 4159659 |
| ME/CFS | Condition | 432738 |
| COVID-19 | Condition | 37311061 |
| Post-acute COVID-19 (Long COVID) | Condition | 705076 |

**Supplementary Table 2: Logic Liaison Conditions** [[31]](https://paperpile.com/c/ycyRlK/bMT8)

| Logic Liaison Table Condition | Codeset IDs |
| --- | --- |
| Obesity | 772561500 |
| Kidney Disease | 1000049286 |
| Pregnancy | 101607593 |
| Rheumatologic Disease | 437151010 |
| Tobacco Smoker | 166730099 |
| Solid Organ Blood Stem Cell Transplant | 225621426 |
| Substance Use Disorder | 1000043391 |
| Long COVID Clinic Visit | 1000032668 |
| Suspected Covid-19 | 229223655 |
| B94.8 | 783054135 |
| Cardiomyopathies | 175678032 |
| Cerebrovascular Disease | 937546319 |
| Chronic Lung Disease | 674884280 |
| Congestive Heart Failure | 1000044269 |
| Coronary Artery Disease | 313241620 |
| Dementia | 963389709 |
| Depression | 963389709 |
| Diabetes Complicated | 263424204 |
| Diabetes Uncomplicated | 689624081 |
| Down Syndrome | 859831250 |
| Heart Failure | 1000038723 |
| Hemiplegia or Paraplegia | 174172884 |
| HIV Infection | 490831143 |
| Hypertension | 211352892 |
| COVID Diagnosis | 1000012899 |
| Long COVID Diagnosis | 1000042623 |
| Multisystem Inflammatory Syndrome | 200799555 |
| Pneumonia due to COVID | 698347441 |
| Malignant Cancer | 1000073908 |
| Metastatic Solid Tumor Cancers | 1000080916 |
| Mild Liver Disease | 17655741 |
| Moderate to Severe Liver Disease | 285109253 |
| Myocardial Infarctions | 944073846 |
| Autoimmune Disease/Immunodeficiency | 1000079426 |
| Peptic Ulcer | 1000089753 |
| Peripheral Vascular Disease | 1000020459 |
| Psychosis | 361905128 |
| Pulmonary Embolism | 503985771 |
| Sickle Cell Disease | 1000028033 |
| Substance Abuse | 606624084 |
| Thalassemia | 1000047917 |
| Tuberculosis | 772103963 |
| Nirmatrelvir | 399252964 |
| Ritonavir | 329050933 |
| Paxlovid | 123908016 |
| Remdesivir | 719693192 |
| N3C Corticosteroids for Systemic Use | 363331988 |
| Antibody Positive | 871267146, 1000005668 |
| Antibody Negative | 871267146, 1000061534 |
| PCR AG Positive | 1000019497, 1000005668 |
| PCR AG Negative | 1000019497, 1000061534 |

**Supplementary Table 3: Covariates of Cases and Controls**

| **Variable** | **hEDS** | **Control** | **Standard Deviation hEDS** | **Standard Deviation Control** |
| --- | --- | --- | --- | --- |
| **Age** | 37.23 | 37.60 | 15.60 | 21.66 |
| **Data partner: 1** | 516 | 488 |  |  |
| **Data partner: 10** | 990 | 1218 |  |  |
| **Data partner: 11** | 39 | 51 |  |  |
| **Data partner: 12** | 647 | 430 |  |  |
| **Data partner: 13** | 54 | 91 |  |  |
| **Data partner: 14** | 359 | 328 |  |  |
| **Data partner: 15** | 287 | 213 |  |  |
| **Data partner: 16** | 413 | 422 |  |  |
| **Data partner: 17** | 511 | 630 |  |  |
| **Data partner: 18** | 237 | 116 |  |  |
| **Data partner: 19** | 1274 | 687 |  |  |
| **Data partner: 2** | 201 | 203 |  |  |
| **Data partner: 20** | 209 | 162 |  |  |
| **Data partner: 21** | 262 | 185 |  |  |
| **Data partner: 22** | 587 | 346 |  |  |
| **Data partner: 23** | 22 | 116 |  |  |
| **Data partner: 24** | 441 | 626 |  |  |
| **Data partner: 25** | <20 | 71 |  |  |
| **Data partner: 26** | 124 | 117 |  |  |
| **Data partner: 27** | 316 | 687 |  |  |
| **Data partner: 28** | 889 | 1149 |  |  |
| **Data partner: 29** | 216 | 179 |  |  |
| **Data partner: 3** | 82 | 47 |  |  |
| **Data partner: 30** | 151 | 107 |  |  |
| **Data partner: 31** | 1426 | 527 |  |  |
| **Data partner: 32** | 471 | 295 |  |  |
| **Data partner: 33** | 849 | 307 |  |  |
| **Data partner: 34** | 2194 | 1537 |  |  |
| **Data partner: 35** | 143 | 195 |  |  |
| **Data partner: 36** | 242 | 342 |  |  |
| **Data partner: 37** | 187 | 238 |  |  |
| **Data partner: 38** | 218 | 346 |  |  |
| **Data partner: 39** | 644 | 1407 |  |  |
| **Data partner: 4** | 41 | 63 |  |  |
| **Data partner: 40** | 572 | 445 |  |  |
| **Data partner: 41** | 1478 | 1532 |  |  |
| **Data partner: 42** | 154 | 154 |  |  |
| **Data partner: 43** | 23 | 36 |  |  |
| **Data partner: 44** | 848 | 637 |  |  |
| **Data partner: 45** | 75 | 96 |  |  |
| **Data partner: 46** | 131 | 175 |  |  |
| **Data partner: 47** | 171 | 561 |  |  |
| **Data partner: 48** | 249 | 502 |  |  |
| **Data partner: 49** | 383 | 307 |  |  |
| **Data partner: 5** | 78 | 313 |  |  |
| **Data partner: 50** | 248 | 205 |  |  |
| **Data partner: 51** | 361 | 190 |  |  |
| **Data partner: 52** | 155 | 101 |  |  |
| **Data partner: 53** | 95 | 145 |  |  |
| **Data partner: 54** | 287 | 182 |  |  |
| **Data partner: 55** | 123 | 173 |  |  |
| **Data partner: 56** | 41 | 410 |  |  |
| **Data partner: 57** | 465 | 539 |  |  |
| **Data partner: 58** | 655 | 731 |  |  |
| **Data partner: 59** | 701 | 688 |  |  |
| **Data partner: 6** | 157 | 204 |  |  |
| **Data partner: 60** | 489 | 385 |  |  |
| **Data partner: 7** | 515 | 511 |  |  |
| **Data partner: 8** | 241 | 70 |  |  |
| **Data partner: 9** | 1209 | 1934 |  |  |
| **ED visits** | 2.93 | 2.73 | 6.85 | 8.91 |
| **Inpatient visits** | 0.99 | 0.98 | 3.30 | 3.70 |
| **Outpatient visits** | 109.46 | 104.42 | 132.66 | 163.51 |
| **Race/Ethnicity: American Indian or Alaska Native Non-Hispanic** | 143 | 163 |  |  |
| **Race/Ethnicity: Asian Non-Hispanic** | 254 | 226 |  |  |
| **Race/Ethnicity: Black or African American Non-Hispanic** | 657 | 611 |  |  |
| **Race/Ethnicity: Hispanic or Latino Any Race** | 1381 | 1181 |  |  |
| **Race/Ethnicity: Native Hawaiian or Other Pacific Islander Non-Hispanic** | 34 | 29 |  |  |
| **Race/Ethnicity: Other Non-Hispanic** | 254 | 200 |  |  |
| **Race/Ethnicity: Unknown** | 1001 | 889 |  |  |
| **Race/Ethnicity: White Non-Hispanic** | 21428 | 21853 |  |  |
| **Race: American Indian or Alaska Native** | 175 | 198 |  |  |
| **Race: Asian** | 257 | 229 |  |  |
| **Race: Black or African American** | 686 | 644 |  |  |
| **Race: Hispanic or Latino** | <20 | <20 |  |  |
| **Race: Native Hawaiian or Other Pacific Islander** | 38 | 32 |  |  |
| **Race: Other** | 349 | 301 |  |  |
| **Race: Unknown** | 1428 | 1257 |  |  |
| **Race: White** | 22218 | 22491 |  |  |
| **Sex: FEMALE** | 21180 | 21607 |  |  |
| **Sex: MALE** | 3214 | 2830 |  |  |
| **Sex: No matching concept** | 749 | 709 |  |  |
| **Sex: UNKNOWN** | <20 | <20 |  |  |
| **Total visits** | 134.01 | 126.68 | 152.35 | 186.26 |

Table 3: Showing raw counts for categorical variables between cases (hEDS) and controls. Continuous variables are averaged, with standard deviation shown. All counts <20 are shown as such for patient protection. Data partner IDs are obfuscated for clinical site protection.

**Supplementary Table 4: Odds ratio values and statistics**

| **condition_name** | **odds_ratio** | **lower_CI** | **upper_CI** | **p_value** | **p_value_FDR** |
| --- | --- | --- | --- | --- | --- |
| **Antibody_Pos_indicator** | 0.69 | 0.66 | 0.71 | 4.00E-71 | 3.47E-70 |
| **Antibody_Neg_indicator** | 1.14 | 1.09 | 1.19 | 7.01E-08 | 1.61E-07 |
| **PCR_AG_Pos_indicator** | 0.69 | 0.66 | 0.71 | 4.00E-71 | 3.47E-70 |
| **TOBACCOSMOKER_indicator** | 1.08 | 1.03 | 1.13 | 7.01E-04 | 0.001333912 |
| **SOLIDORGANORBLOODSTEMCELLTRANSPLANT_indicator** | 0.57 | 0.49 | 0.65 | 2.09E-16 | 7.42E-16 |
| **SUBSTANCEUSEDISORDER_indicator** | 1.16 | 1.11 | 1.21 | 3.41E-12 | 9.49E-12 |
| **MODERATESEVERELIVERDISEASE_indicator** | 0.48 | 0.41 | 0.57 | 2.68E-17 | 1.10E-16 |
| **MYOCARDIALINFARCTION_indicator** | 0.64 | 0.57 | 0.71 | 4.69E-17 | 1.74E-16 |
| **OTHERIMMUNOCOMPROMISED_indicator** | 2.05 | 1.94 | 2.15 | 3.36E-168 | 6.56E-167 |
| **DOWNSYNDROME_indicator** | 0.38 | 0.23 | 0.62 | 1.36E-04 | 2.66E-04 |
| **HEARTFAILURE_indicator** | 0.60 | 0.55 | 0.65 | 3.15E-38 | 1.89E-37 |
| **HEMIPLEGIAORPARAPLEGIA_indicator** | 1.04 | 0.90 | 1.19 | 0.62066735 | 0.880219151 |
| **CORONARYARTERYDISEASE_indicator** | 0.62 | 0.57 | 0.67 | 3.62E-34 | 2.02E-33 |
| **DEMENTIA_indicator** | 0.46 | 0.37 | 0.56 | 4.93E-14 | 1.54E-13 |
| **DEPRESSION_indicator** | 2.23 | 2.15 | 2.32 | 7.23E-24 | 4.38E-23 |
| **B94_8_indicator** | 2.41 | 1.90 | 3.05 | 4.24E-13 | 1.22E-12 |
| **CARDIOMYOPATHIES_indicator** | 0.66 | 0.59 | 0.74 | 8.18E-14 | 2.45E-13 |
| **CEREBROVASCULARDISEASE_indicator** | 1.11 | 1.03 | 1.20 | 0.005526243 | 0.009796521 |
| **PSYCHOSIS_indicator** | 1.09 | 0.96 | 1.23 | 0.196440821 | 0.306447681 |
| **PULMONARYEMBOLISM_indicator** | 1.04 | 0.92 | 1.18 | 0.500061018 | 0.735938856 |
| **SICKLECELLDISEASE_indicator** | 0.53 | 0.29 | 0.96 | 0.03683679 | 0.062462384 |
| **LL_MISC_indicator** | 0.69 | 0.20 | 2.44 | 0.566103686 | 0.817705324 |
| **LL_PNEUMONIADUETOCOVID_indicator** | 0.66 | 0.56 | 0.78 | 9.54E-07 | 2.13E-06 |
| **MALIGNANTCANCER_indicator** | 0.67 | 0.63 | 0.72 | 5.43E-34 | 2.82E-33 |
| **METASTATICSOLIDTUMORCANCERS_indicator** | 0.18 | 0.08 | 0.38 | 7.02E-06 | 1.48E-05 |
| **MILDLIVERDISEASE_indicator** | 1.04 | 0.98 | 1.11 | 0.156213894 | 0.253847578 |
| **LL_Long_COVID_clinic_visit_indicator** | 2.98 | 2.18 | 4.08 | 7.36E-12 | 1.98E-11 |
| **LL_SUSPECTEDCOVID19_indicator** | 0.78 | 0.56 | 1.11 | 0.165595327 | 0.263600725 |
| **PAX1_NIRMATRELVIR_indicator** | 1.09 | 0.92 | 1.28 | 0.312454918 | 0.468682377 |
| **PAX2_RITONAVIR_indicator** | 1.10 | 0.94 | 1.30 | 0.232402901 | 0.355439731 |
| **PAXLOVID_indicator** | 2.05 | 1.89 | 2.22 | 5.43E-70 | 4.24E-69 |
| **confirmed_covid_patient** | 0.89 | 0.86 | 0.92 | 2.52E-10 | 6.55E-10 |
| **possible_covid_patient** | 2.93 | 2.44 | 3.51 | 3.28E-31 | 1.50E-30 |
| **patient_death_indicator** | 0.43 | 0.37 | 0.49 | 3.29E-32 | 1.60E-31 |
| **CHRONICLUNGDISEASE_indicator** | 1.96 | 1.88 | 2.04 | 1.55E-233 | 4.03E-232 |
| **CONGESTIVEHEARTFAILURE_indicator** | 0.42 | 0.38 | 0.46 | 3.01E-63 | 1.96E-62 |
| **PEPTICULCER_indicator** | 1.54 | 1.39 | 1.70 | 3.28E-17 | 1.28E-16 |
| **PERIPHERALVASCULARDISEASE_indicator** | 0.75 | 0.68 | 0.82 | 3.20E-09 | 7.79E-09 |
| **REMDISIVIR_indicator** | 0.63 | 0.53 | 0.76 | 1.48E-06 | 3.21E-06 |
| **SYSTEMICCORTICOSTEROIDS_indicator** | 2.10 | 2.02 | 2.19 | 1.38E-289 | 5.38E-288 |
| **LL_COVID_diagnosis_indicator** | 1.06 | 1.02 | 1.11 | 0.001947966 | 0.00353352 |
| **LL_Long_COVID_diagnosis_indicator** | 2.88 | 2.55 | 3.26 | 2.57E-64 | 1.82E-63 |
| **DIABETESCOMPLICATED_indicator** | 0.49 | 0.46 | 0.53 | 7.63E-82 | 9.92E-81 |
| **DIABETESUNCOMPLICATED_indicator** | 0.59 | 0.55 | 0.62 | 3.44E-76 | 3.83E-75 |
| **PREGNANCY_indicator** | 0.89 | 0.85 | 0.94 | 4.79E-05 | 9.57E-05 |
| **RHEUMATOLOGICDISEASE_indicator** | 2.09 | 1.97 | 2.21 | 1.90E-134 | 2.96E-133 |
| **SUBSTANCEABUSE_indicator** | 0.88 | 0.81 | 0.95 | 0.001144735 | 0.002125936 |
| **THALASSEMIA_indicator** | 1.63 | 1.02 | 2.61 | 0.041400011 | 0.068706401 |
| **TUBERCULOSIS_indicator** | 5.20 | 3.69 | 7.33 | 3.97E-21 | 1.72E-20 |
| **MOLNUPIRAVIR_indicator** | 1.48 | 1.23 | 1.79 | 4.43E-05 | 9.09E-05 |
| **HIVINFECTION_indicator** | 0.71 | 0.51 | 0.98 | 0.036257001 | 0.062462384 |
| **HYPERTENSION_indicator** | 0.84 | 0.81 | 0.88 | 2.74E-15 | 9.30E-15 |
| **OBESITY_indicator** | 1.12 | 1.08 | 1.16 | 2.15E-09 | 5.40E-09 |
| **KIDNEYDISEASE_indicator** | 0.78 | 0.74 | 0.84 | 3.90E-14 | 1.27E-13 |
| **PCR_AG_Neg_indicator** | 1.14 | 1.09 | 1.19 | 7.01E-08 | 1.61E-07 |

Table 4. This table presents the odds ratios (ORs) and 95% confidence intervals (CIs) for each condition comparing patients with hEDS to matched controls. FDR-adjusted q-values are also provided to account for multiple testing across all conditions. ORs with q < 0.05 are considered statistically significant after false discovery rate correction. Analyses were performed using conditional logistic regression accounting for matched pairs and healthcare utilization.

**Supplementary Table 5: Prevalence Rates of Comorbidities Segmented by Long COVID Flag**

| **eds_flag** | **long_covid_flag** | **mcas_prevalence** | **pots_prevalence** | **mecfs_prevalence** | **mcas_pots_prevalence** | **mcas_mecfs_prevalence** | **pots_mecfs_prevalence** | **all_three_prevalence** |
| --- | --- | --- | --- | --- | --- | --- | --- | --- |
| **1** | 0 | 9.5 | 21.7 | 14.6 | 5.3 | 2.8 | 5.2 | 1.7 |
| **1** |  | <20 | <20 | <20 | <20 | <20 | <20 | <20 |
| **1** | 1 | 25.3 | 42.7 | 42.0 | 19.4 | 14.1 | 21.8 | 11.9 |
| **0** | 0 | <20 | <20 | 3.3 | <20 | <20 | <20 | <20 |
| **0** | 1 | <20 | <20 | 22.84 | <20 | <20 | <20 | <20 |
| **0** |  | <20 | <20 | <20 | <20 | <20 | <20 | <20 |

Table 5. This table shows the prevalence of individual comorbidities and their combinations among patients stratified by hEDS diagnosis (EDS flag) and Long COVID status (Long COVID flag). Prevalence is reported as the proportion of patients with each condition within the respective subgroup. Combinations of comorbidities are included to highlight patterns of co-occurrence. These data provide context for phenotypic clustering and potential interactions between hEDS and Long COVID.

**Supplemental Table 6: Cumulative incidence of hEDS diagnoses by comorbidity group and index condition.**

| **Index Condition** | **Comorbidity Group** | **Day 180 Cum. Incidence (%)** | **Day 365 Cum. Incidence (%)** | **Day 730 Cum. Incidence (%)** | **Log-rank p vs hEDS+POTS** | **Log-rank p vs hEDS+MCAS** | **Log-rank p vs hEDS+ME/CFS** | **Log-rank p vs hEDS** |
| --- | --- | --- | --- | --- | --- | --- | --- | --- |
| COVID | hEDS+POTS | 75.46 | 58.48 | 29.98 | - | 0.1179 | 0.0000 | 0.0000 |
| COVID | hEDS+MCAS | 77.19 | 55.85 | 24.36 | 0.1179 | - | 0.0000 | 0.0000 |
| COVID | hEDS+ME/CFS | 88.78 | 79.15 | 61.15 | 0.0000 | 0.0000 | - | 0.0000 |
| COVID | hEDS | 94.64 | 91.32 | 85.72 | 0.0000 | 0.0000 | 0.0000 | - |
| Long COVID | hEDS+POTS | 78.73 | 61.58 | 38.22 | - | 0.1643 | 0.0005 | 0.0000 |
| Long COVID | hEDS+MCAS | 77.78 | 55.12 | 30.85 | 0.1643 | - | 0.0000 | 0.0000 |
| Long COVID | hEDS+ME/CFS | 85.34 | 74.00 | 58.09 | 0.0005 | 0.0000 | - | 0.0000 |
| Long COVID | hEDS | 92.35 | 88.65 | 84.80 | 0.0000 | 0.0000 | 0.0000 | - |

This table presents cumulative incidence (%) of hEDS diagnoses at 180, 365, and 730 days for patients stratified by comorbidity group (POTS, MCAS, ME/CFS, or none) and index condition (COVID-19 or Long COVID). Log-rank p-values are shown for pairwise comparisons between each comorbidity group and reference groups. A dash (“–”) indicates comparison with the same group. These data provide insight into the temporal patterns of hEDS diagnosis across comorbidities and infection status.

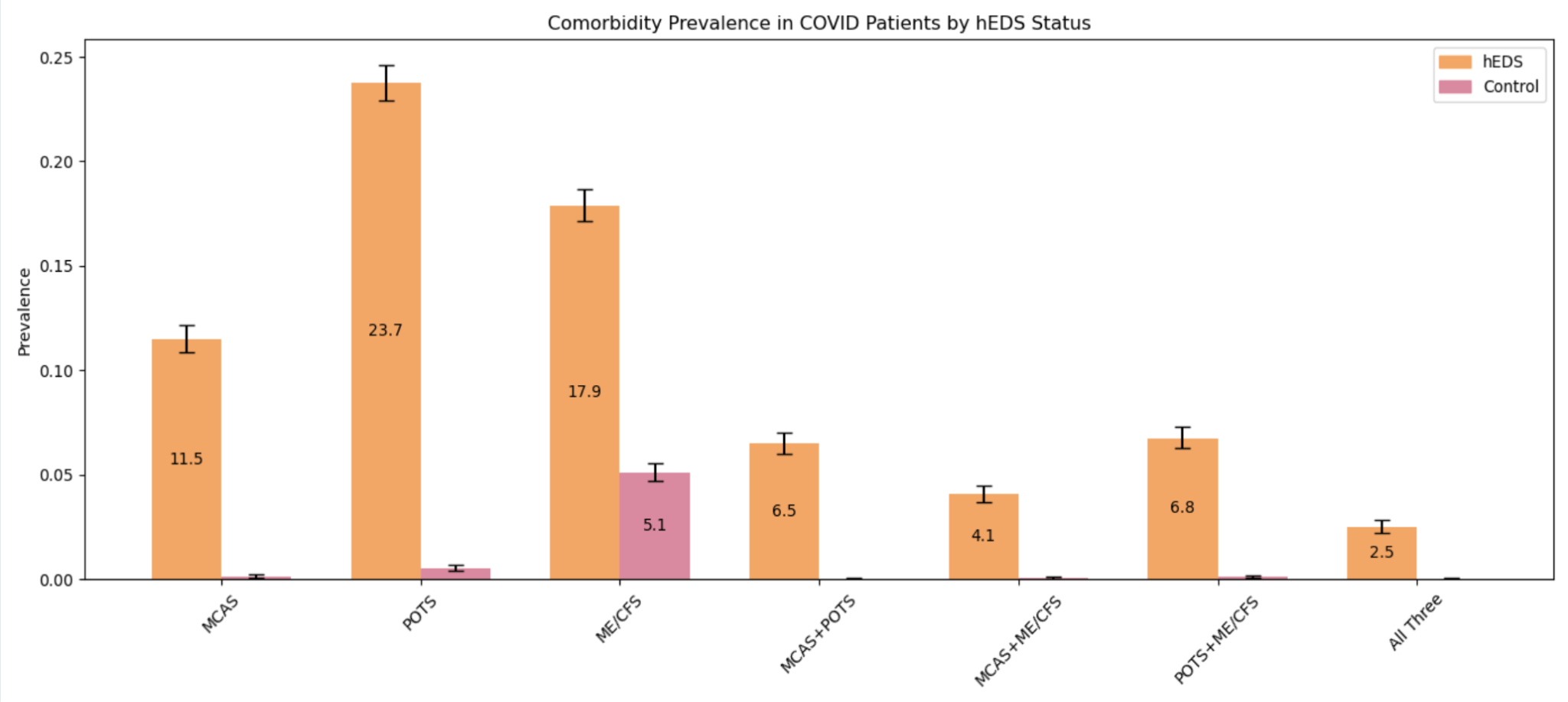

**Supplemental Figure 1. Prevalence of comorbid conditions in COVID-19 patients by hEDS status.**

Bar heights represent the prevalence of commonly reported comorbid conditions among patients with hEDS and COVID-19 (n = 25,152) versus matched controls with COVID-19 (n = 25,152). Prevalence for combinations of conditions is also shown. To protect patient confidentiality, counts fewer than 20 are reported as <20 or left blank. Overall, patients with hEDS exhibit higher prevalence of these comorbidities, suggesting that pre-existing conditions may contribute to risk of complications following COVID-19.

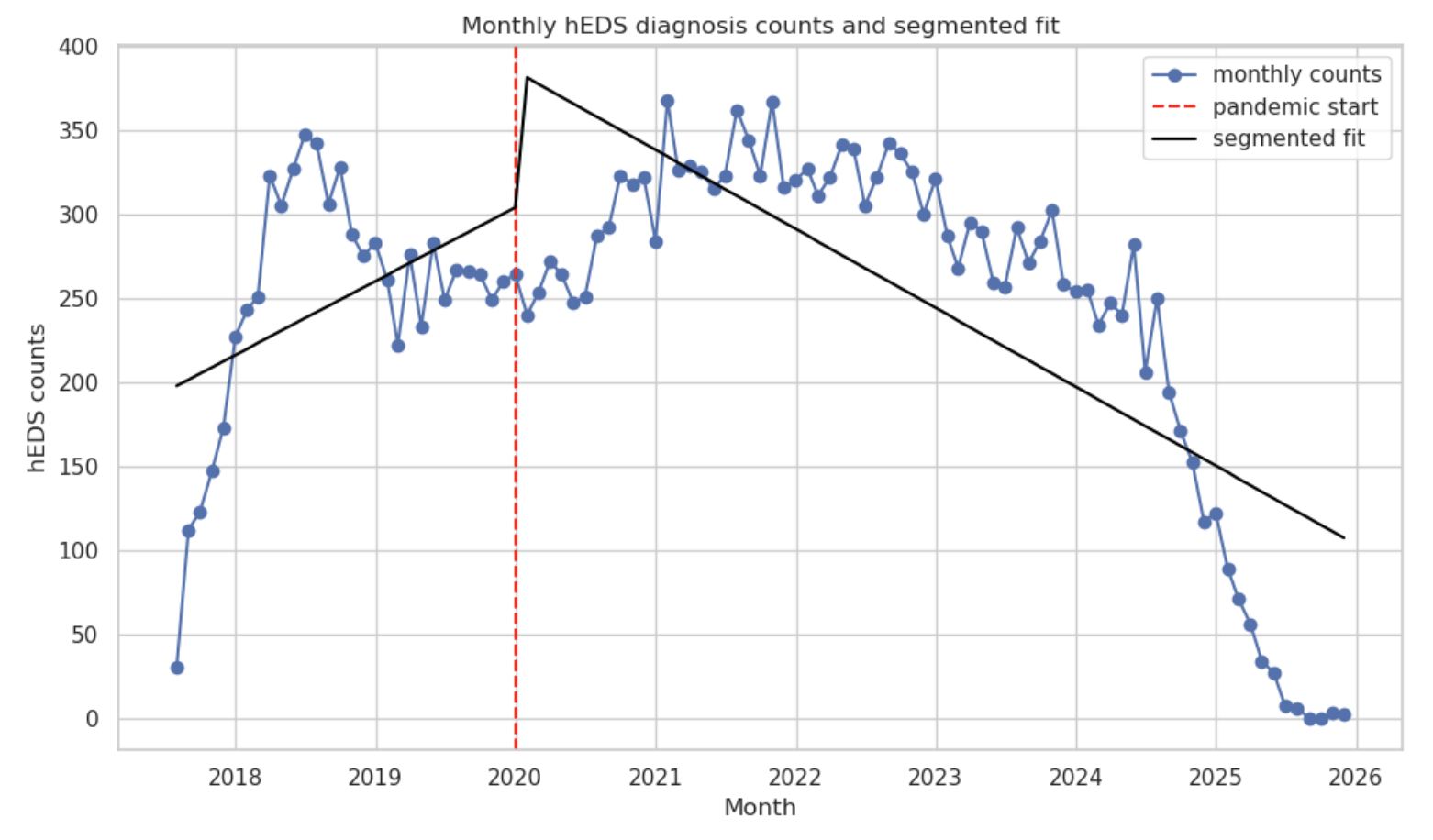

**Supplemental Figure 2. Interrupted time series of monthly hEDS diagnosis counts (2018–2026).**
 Monthly counts of hEDS diagnoses are shown from January 2018 through 2026. The vertical dashed line indicates the start of the COVID-19 pandemic (January 2020), and segmented regression lines illustrate the trend before and after this interruption. This analysis allows visualization of temporal changes in hEDS diagnoses over time and the potential impact of the pandemic on diagnostic patterns.

**Supplemental Figure 3: Cumulative incidence plots show patients with hEDS have higher likelihood of receiving a diagnosis after an acute post viral infection with complications when comorbidities are present.**

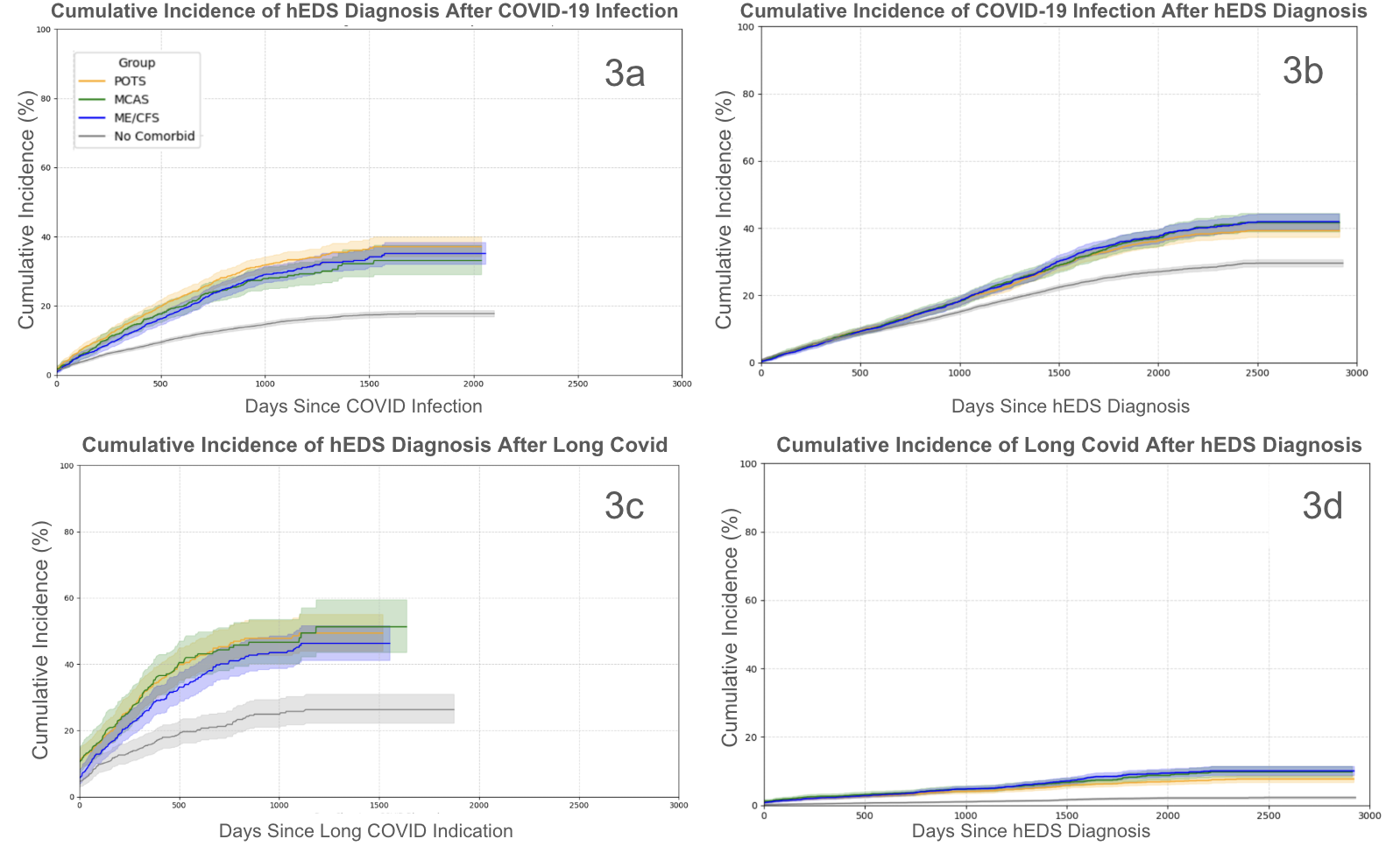

**Supplemental Figure 3: Cumulative incidence plots show patients with hEDS have higher likelihood of receiving a diagnosis after an acute post viral infection with complications when comorbidities are present.** Cumulative incidence plots demonstrate that patients with hEDS have a higher likelihood of receiving a diagnosis following acute post-viral infection and experience greater risk of complications when comorbid conditions are present. Panels **3a and c** show the cumulative incidence of receiving an hEDS diagnosis after COVID-19 infection (**3a)** or Long COVID (**3c)**. Panels **3b and d** assess the cumulative incidence of developing either a COVID-19 infection (**3b)** or Long COVID (**3d)** after receiving an hEDS diagnosis. Time is measured in days from the index event (e.g., COVID-19 diagnosis or hEDS diagnosis), and cumulative incidence was estimated using the Kaplan-Meier method. Shaded regions represent 95% confidence intervals.

| 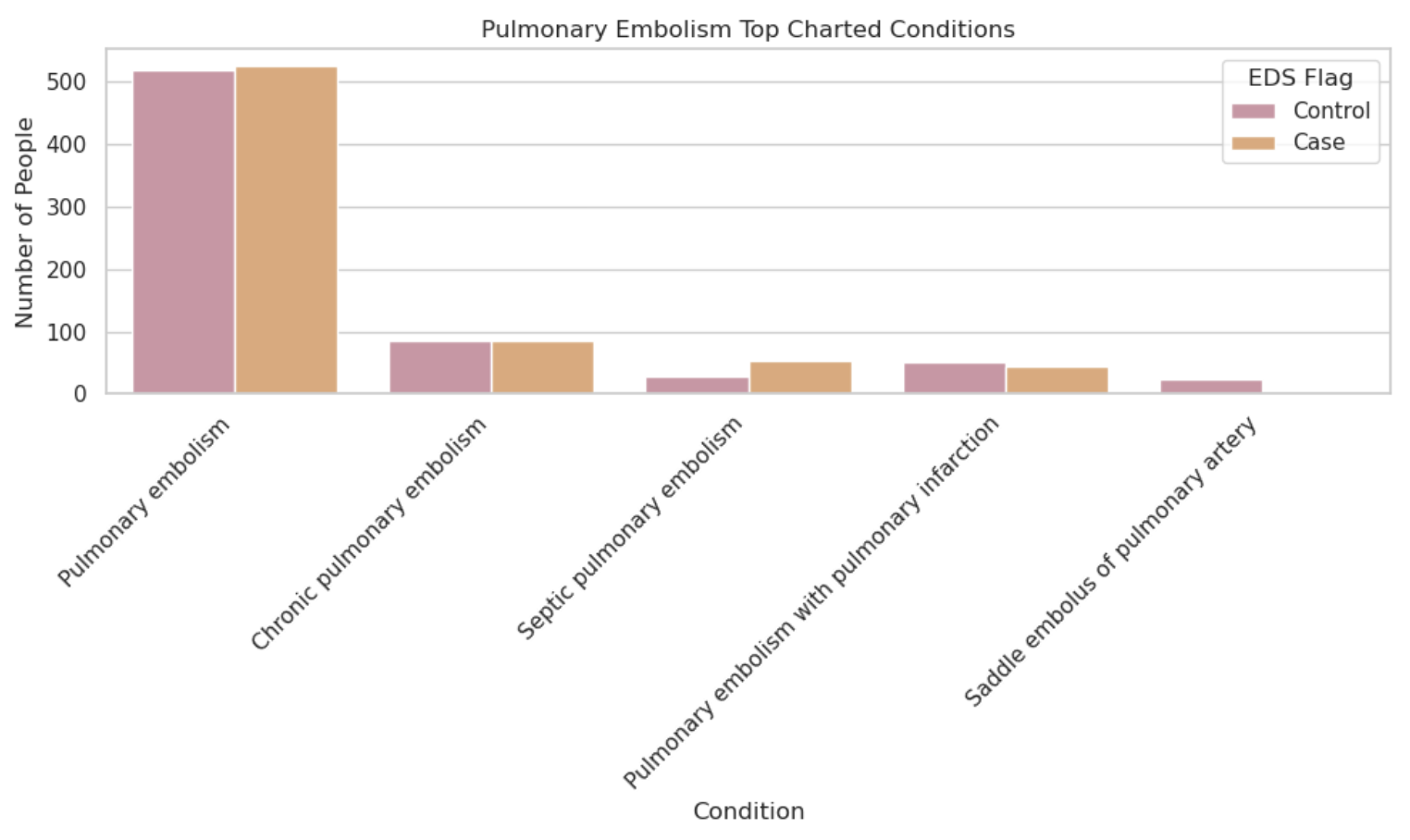**4a** | 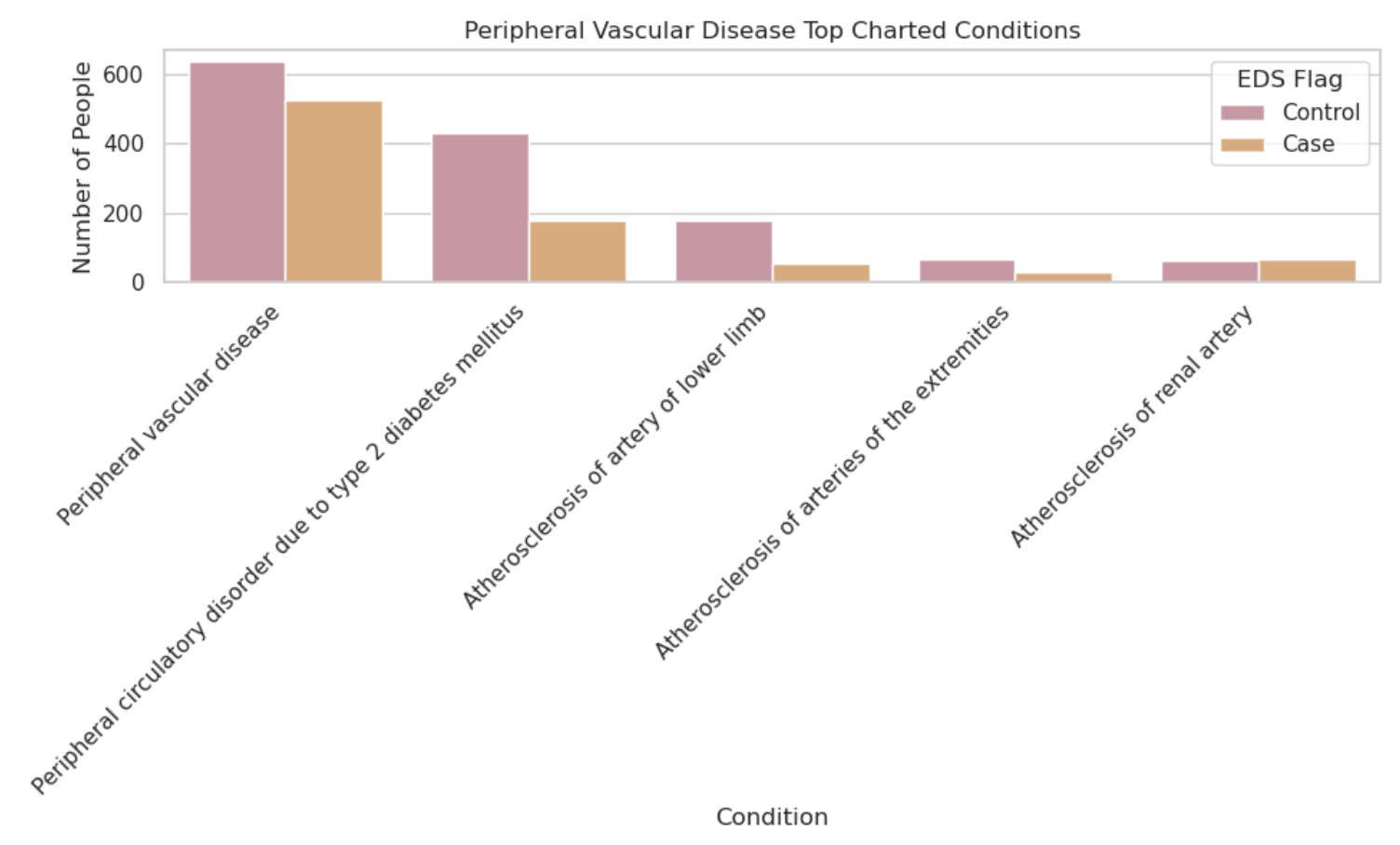**4b** |
| --- | --- |
| 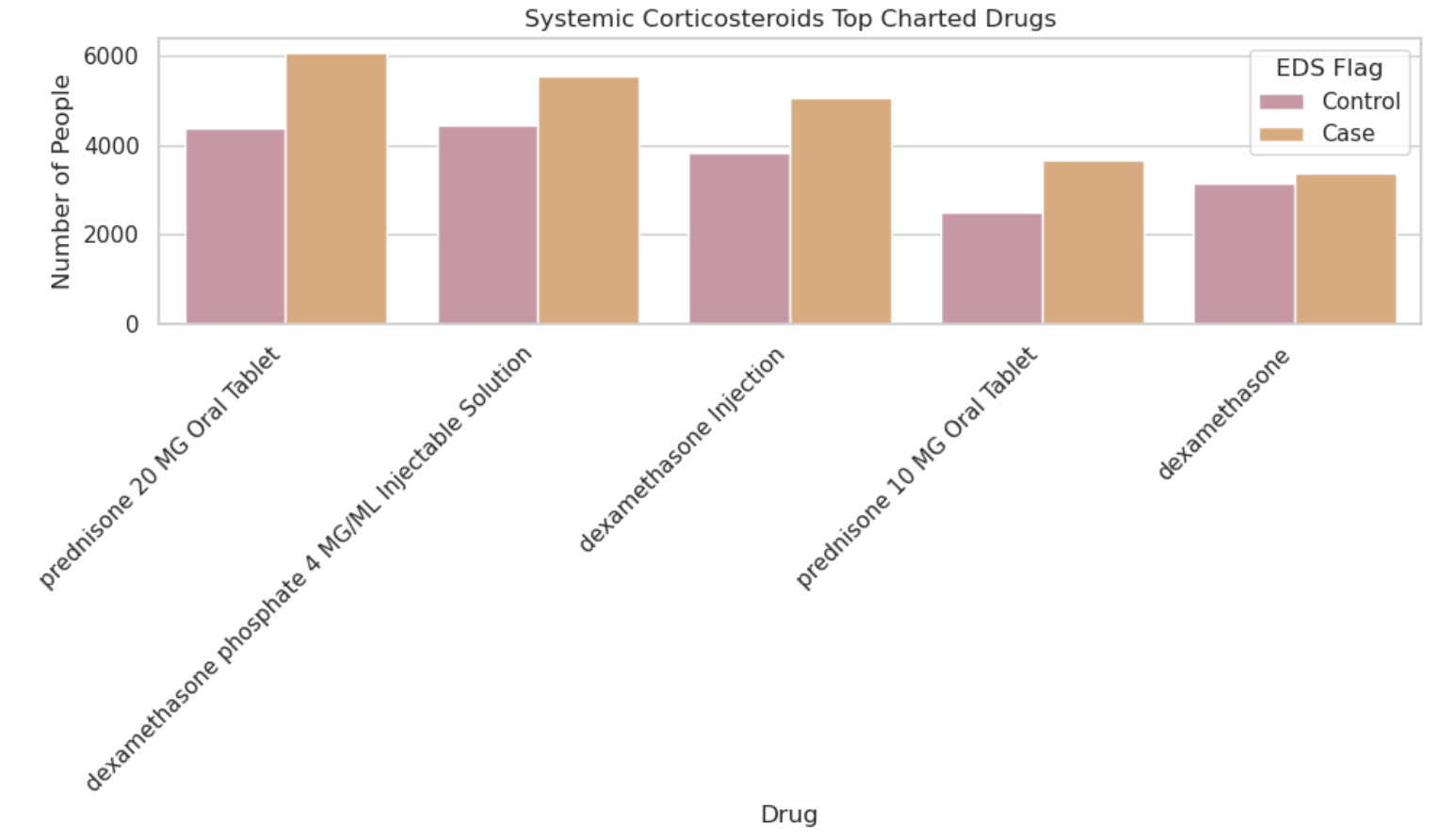**4c** | 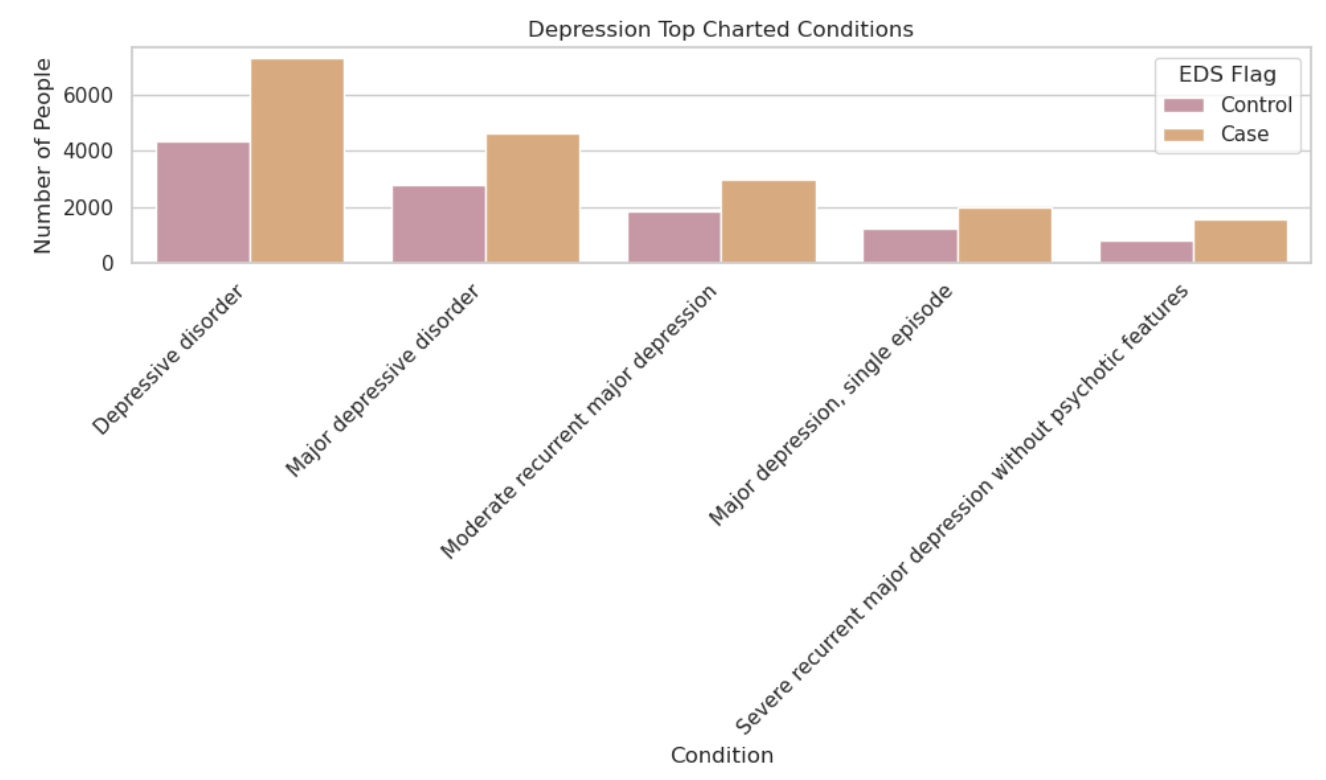**4d** |
| 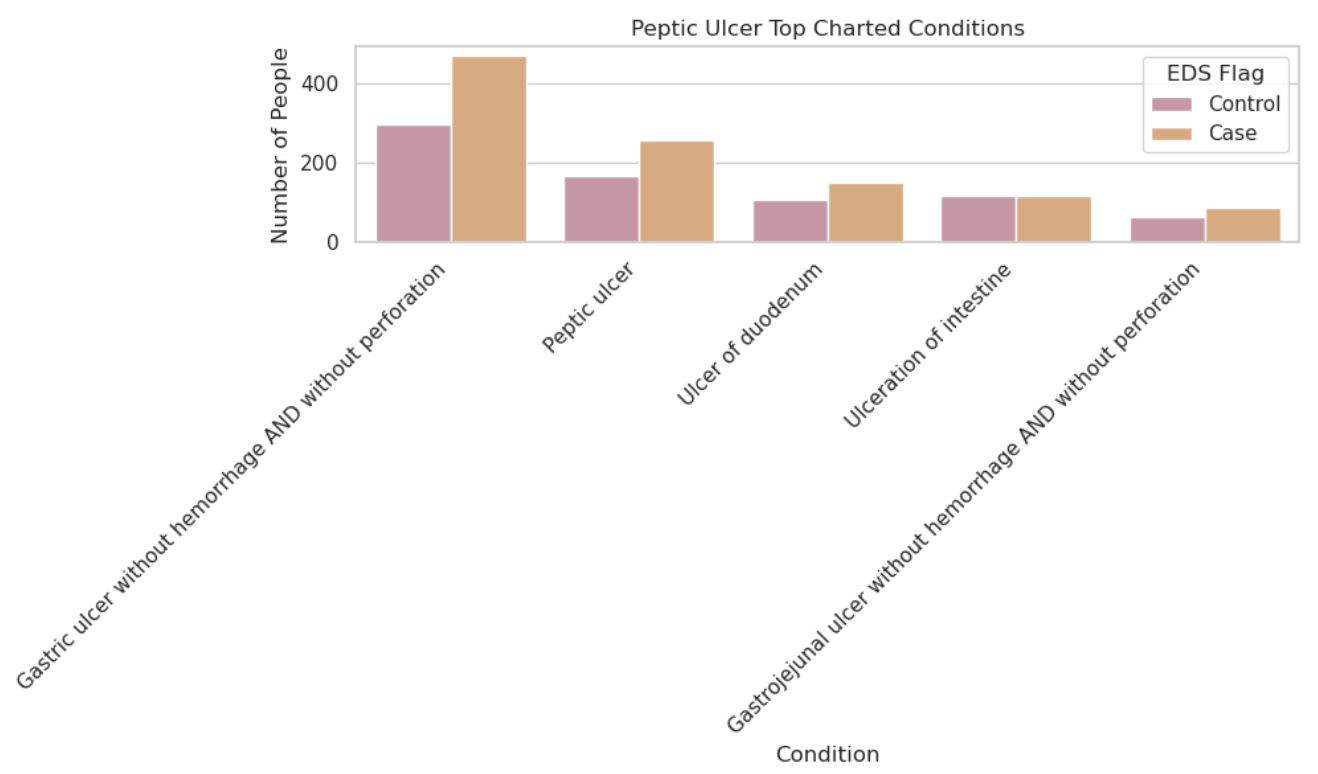**4e** | 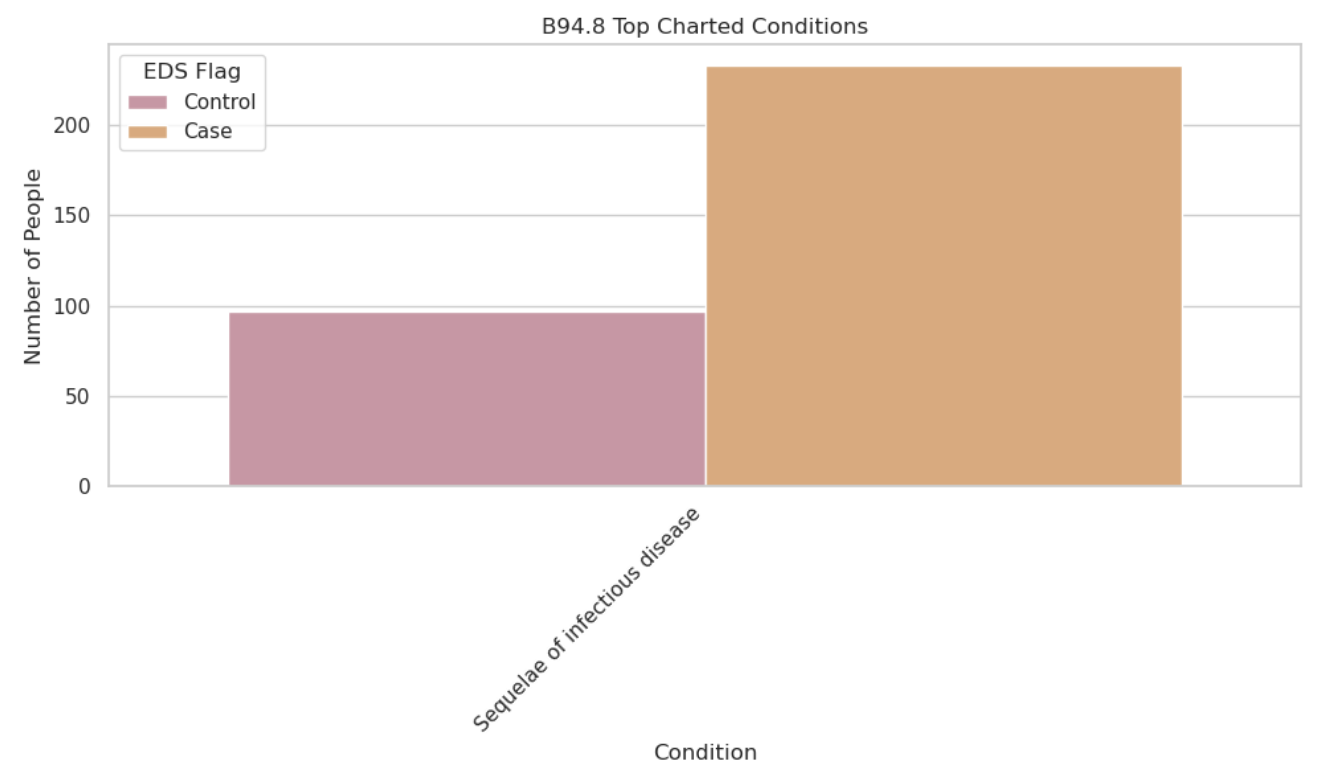**4f** |
| **Supplemental Figure 4. Additional Top Charted Conditions.** Raw counts of top contributing diagnoses of remaining categories are shown: **(4a)** pulmonary embolism, **(4b)** peripheral vascular disease **(4c)** systemic corticosteroids **(4d)** depression, **(4e)** peptic ulcer, and **(4f)** B94.8. All patient counts fewer than 20 are reported as "<20" to protect confidentiality. | |

### **Supplemental Methods: Prevalence Calculation**

Point prevalence rates were calculated to quantify the burden of hypermobile Ehlers-Danlos syndrome (hEDS), associated comorbidities, COVID-19 infection, and Long COVID within the study cohort.

The denominator for all prevalence estimates included all individuals in the N3C Enclave with at least one clinical encounter between January 1, 2018, and May 1, 2025, who also had complete demographic data (sex, age, race, and ethnicity). The numerators were defined by the number of individuals meeting case criteria for hEDS, each comorbidity, COVID-19, or Long COVID, based on OMOP concept sets as described in the cohort construction and comorbidity definitions.

This approach captures the proportion of patients diagnosed with the condition at any point during the study period, regardless of their current status.
